# Modelling to inform the COVID-19 response in Bangladesh

**DOI:** 10.1101/2021.04.19.21255673

**Authors:** Elaine A Ferguson, Eric Brum, Anir Chowdhury, Shayan Chowdhury, Mikolaj Kundegorski, Ayesha S Mahmud, Nabila Purno, Ayesha Sania, Rachel Steenson, Motahara Tasneem, Katie Hampson

## Abstract

**Background:** Non-pharmaceutical interventions (NPIs) used to limit SARS-CoV-2 transmission vary in their feasibility, appropriateness and effectiveness in different contexts. In Bangladesh a national lockdown implemented in March 2020 exacerbated poverty and was untenable long-term, and a resurgence in 2021 warranted renewed NPIs.

**Methods:** We developed an SEIR model for Dhaka District, parameterised from literature values and calibrated to data from Bangladesh. We discussed scenarios and parameterisations with policymakers with the aid of an interactive app. These discussions guided modelling of lockdown and two post-lockdown measures considered feasible to deliver; symptoms-based household quarantining and compulsory mask-wearing. We examined how testing capacity affects case detection, and compared NPI scenarios on deaths, hospitalisations relative to capacity, working days lost, and cost-effectiveness.

**Results:** Lockdowns alone were predicted to delay the first epidemic peak but could not prevent overwhelming of the health service and were costly in lost working days. Impacts of post-lockdown interventions depended heavily on compliance. Assuming 80% compliance, symptoms-based household quarantining alone could not prevent hospitalisations exceeding capacity, whilst mask-wearing prevented overwhelming health services and was cost-effective given masks of high filtration efficiency. Combining masks with quarantine increased their impact. Even at maximum testing capacity, confirmed cases far underestimate total cases. Recalibration to surging cases in 2021 suggests potential for another wave later in 2021, dependent on uncertainties in case reporting and immunity.

**Conclusions:** Masks and symptoms-based household quarantining synergistically prevent transmission, and are cost-effective in Bangladesh. Our interactive app was valuable in supporting decision-making, with mask-wearing being mandated early, and community teams being deployed to support quarantining across Dhaka. These measures likely contributed to averting the worst public health impacts, but delivering an effective response at scale has been challenging. Messaging to increase compliance with mask-wearing and quarantine is needed to reduce the risk and impacts from another wave.

## Introduction

Human infections with severe acute respiratory syndrome coronavirus 2 (SARS-CoV-2), the virus that causes the respiratory disease COVID-19, were first observed in China in December 2019 (Li et al., 2020). By the end of 2020, more than 80.3 million cases and 1.77 million associated deaths had been reported worldwide (European Centre for Disease Prevention and Control, 2021). With limited treatment options, countries turned to a range of non-pharmaceutical interventions (NPIs) to limit transmission. These measures include improved hygiene practices, social distancing, contact tracing, travel restrictions, quarantines, shielding of the vulnerable, lockdowns of differing severity, and facemasks. NPIs have been used by high-income countries (HICs) and low- and middle-income countries (LMICs) alike, but for a variety of reasons some of these measures may be less effective or more difficult to maintain in LMICs. Vaccines have come into play in 2021, but rollout has been almost exclusively in HICs. With negligible vaccination coverage and only NPIs to mitigate impacts, LMICs are now facing subsequent epidemic waves of faster-spreading variants and may have little protective immunity from prior infections (Cele et al., 2021; Planas et al., 2021).

Lockdowns can control COVID-19 epidemics by reducing the effective reproduction number, *R*_*e*_, to below one (Flaxman et al., 2020). However, where many people live precariously without social support, lockdowns can exacerbate poverty (Amewu et al., 2020; Andam et al., 2020) and risk food security, whilst poor adherence limits their effectiveness. Social distancing may be impractical in densely populated areas (Anwar et al., 2020; Chowdhury et al., 2020; Gupta et al., 2020), hindered by larger household sizes and/or cramped conditions (Davies et al., 2020a). Urban slums and refugee camps, where rapid transmission and poor healthcare access co-occur, present a particular concern (Ahmed et al., 2020; Truelove et al., 2020). Shielding the most vulnerable, primarily older people and those with underlying health conditions (Clark et al., 2020), is also challenging in multi-generational households (Hodgins and Saad, 2020; Lloyd-Sherlock et al., 2020; United Nations, Department of Economic and Social Affairs, 2019a). Contact tracing is limited by testing capabilities (facilities, trained personnel, consumables, reagents and biosafety) (Anwar et al., 2020; Homaira et al., 2020; Rahaman et al., 2020) and information management capacity. Moreover, in settings where healthcare resources are already stretched, surge capacity is also low (Torres-Rueda et al., 2020).

Some features of LMICs may mitigate the health costs of any lowered efficiency of NPIs. The relatively younger populations in LMICs, with fewer underlying risk factors, means that a smaller proportion of cases are likely to be severe relative to HICs (Clark et al., 2020; Gupta et al., 2020; Hodgins and Saad, 2020). There has been speculation that the BCG (Bacillus Calmette-Guérin) vaccine, which is no longer routinely used in HICs, but is widely used in LMICs, may confer some protection from severe COVID-19, but evidence from trials is needed to confirm any association (Pereira et al., 2020). More generally, the wider social and economic consequences of NPIs may trade off against their impact on controlling disease, and the structure and underlying health of populations may impact the shape of this trade-off (Reidpath et al., 2020). Hence, there is an urgent need for the development and implementation of contextually appropriate interventions that take into account the population that they target and their needs (Hodgins and Saad, 2020).

During the pandemic, epidemiological models have received increased attention from governments and the public alike, and have influenced policy decisions worldwide (McBryde et al., 2020). However, a translational gap persists between policymakers and scientists working on models for disease forecasting and assessment of interventions. Early in the pandemic, this gap was exacerbated by uncertainties about the biology and transmission of SARS-CoV-2, and the limited information about its spread, given local testing and reporting capacities, not to mention people’s behaviour. As a consequence, there is often simultaneously both overconfidence in, and mistrust of, models.

Ideally, policymakers, scientists and communities should work together to develop and implement locally appropriate interventions. Models can play an important and overlooked role in facilitating this process. By empowering decision-makers to better understand the mechanisms underpinning the timescales and magnitude over which interventions lead to impact, as well as the uncertainties and social and behavioural factors that affect their efficacy, co-created models can inform short- and longer-term policies.

We investigate the potential impact of contextually appropriate NPI strategies to mitigate COVID-19 in Dhaka District, Bangladesh. High-density urban centres and refugee camps in Bangladesh led to fears that strained health systems would be quickly overwhelmed (Anwar et al., 2020). Cases of COVID-19 were first confirmed in Bangladesh on the 8^th^ March 2020, and NPIs were subsequently introduced, beginning with postponements to mass gatherings, followed by international travel restrictions, and culminating in a national lockdown (announced as a ‘general holiday’) from 26^th^ March (Ahmed et al., 2020; Anwar et al., 2020). Rapid movement out of urban areas, particularly from the capital city Dhaka, following the lockdown announcement led to virus spread across the country (Anwar et al., 2020). The lockdown had swift economic repercussions: around 60% of households lost their main income source, the majority of previously vulnerable non-poor households slipped below the poverty line, and food security declined (Rahman et al., 2020). The nationwide lockdown was quickly recognized as untenable long-term (ultimately ending on 1^st^ June), and considered only as a stop-gap measure.

With input from policymakers, we developed an SEIR model to compare the potential of NPIs following the relaxation of lockdown. We designed an associated interactive app within which policymakers could explore these scenarios to gain an understanding of how the model worked and how uncertainties may influence outcomes. Here we compare scenarios based on projected 2020 COVID-19 deaths, and whether hospitalisations remain below capacity. We also explore associated costs and the importance of having NPIs in place by the time lockdown ends. Finally, we examine scenarios following resurgence of cases and a renewed lockdown in March 2021 and update our app for ongoing policy use.

## Methods

### Model description

We developed a deterministic SEIR model comprising a set of ordinary differential equations (ODEs) to describe SARS-CoV-2 transmission in Dhaka District, the most densely populated district in Bangladesh. Dhaka District includes rural areas in addition to the city area itself, but, as case data were resolved to district level, we consider the full district population, rather than restricting analyses to the city population.

The model includes three infectious states, with latently infected individuals either becoming pre-symptomatically infectious (*I*_*p*_) before progressing to symptomatic infection (*I*_*s*_), or becoming asymptomatically infectious (*I*_*a*_) until their recovery (Fig. 1). Susceptible individuals are exposed to SARS-CoV-2 according to transmission rates specific to each infectious state (*β*^*a*^, *β*^*p*^ and *β*^*s*^). These rates are based on: 1) asymptomatic individuals producing 65% of the secondary infections produced by pre-symptomatic-to-symptomatic individuals (Yi et al., 2020); 2) 35% of secondary infections from pre-symptomatic-to-symptomatic individuals happening in the pre-symptomatic period (Liu et al., 2020); and 3) the value of *R*_0_ :

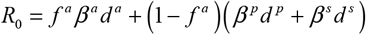

where *d*^*a*^ = 7, *d*^*p*^ = 2 and *d*^*s*^ = 7 are the mean durations (in days) of each infectious state (Byrne et al., 2020; Hu et al., 2020), and *f*^*a*^ is the proportion of infections that are asymptomatic (supplementary Table S4). Recovered individuals are considered to be immune.

**Figure 1:**
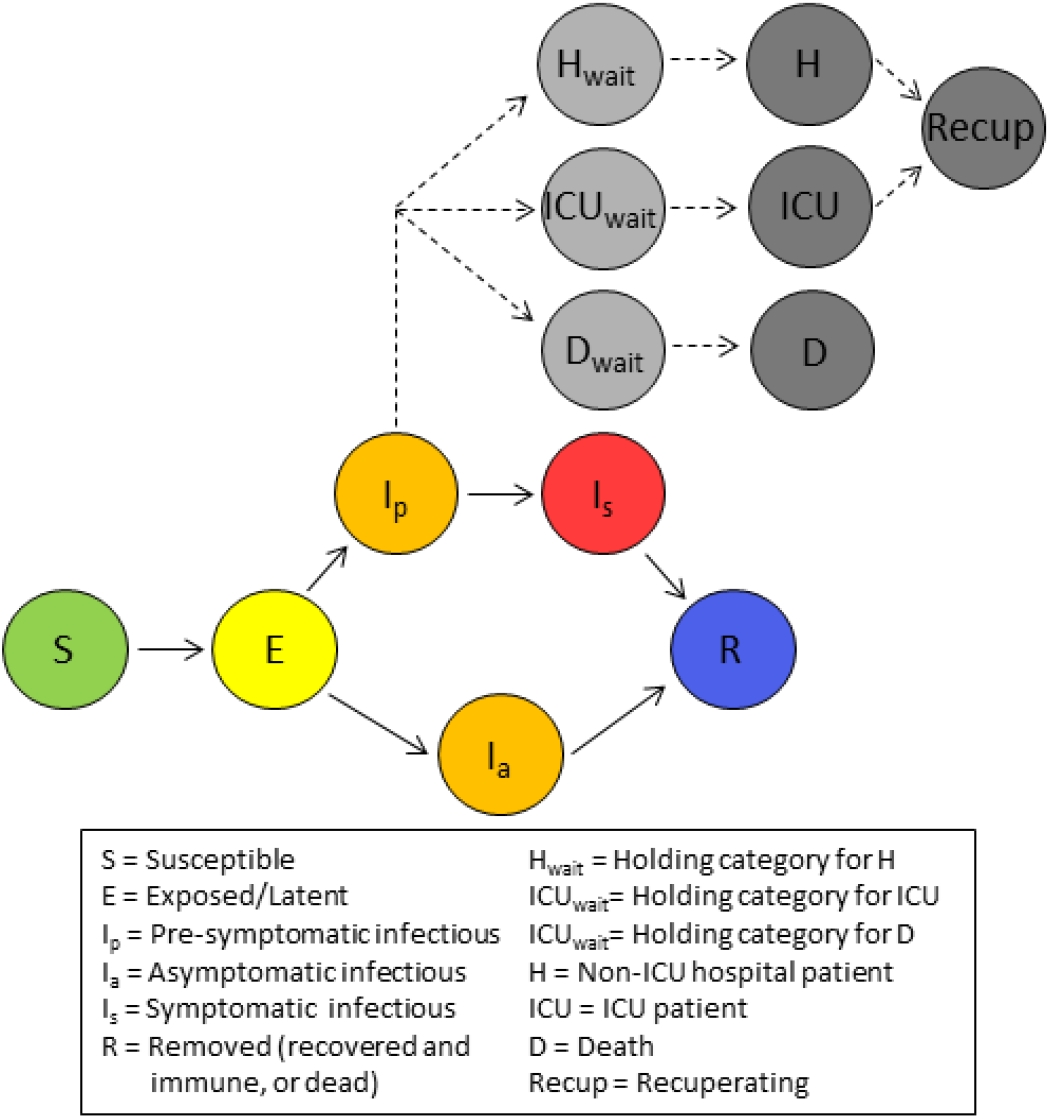
Model Schematic illustrating the movement of individuals between classes. Coloured circles indicate disease states that impact transmission. Grey circles describe health outcomes that are tracked but have no impact on disease transmission.

We track numbers of both general and Intensive Care Unit (ICU) beds required by COVID-19 patients, and deaths due to COVID-19 (Fig. 1). A proportion of individuals *f*^*H*^ (1 - *f*^*ICU*^) that leave the pre-symptomatic state join a holding category prior to progressing to general hospital beds, while a second proportion *f*^*H*^ *f*^*ICU*^ enters a holding category prior to entering ICU beds. Individuals remain in these two holding categories from the first appearance of symptoms until the point at which hospitalisation is required, on average 7 days (Liang et al., 2020; Linton et al., 2020). A third holding category retains a proportion of individuals *f*^*D*^ from the first appearance of symptoms until death, on average 20.2 days later (Linton et al., 2020; Verity et al., 2020). While we assume no overlap between the groups in ICU versus general hospital beds, overlap between those that die and the hospitalized groups is both permitted and expected. On leaving hospital, after a mean of 5 days for general beds and 7 days for ICU (Rees et al., 2020), recovered individuals recuperate for an average of 3 weeks before resuming work, if employed (Halpin et al., 2020). The number of individuals in the seven health outcome states has no impact on transmission dynamics, and it is assumed that there is no impact of a lack of hospital beds on the COVID-19 death rate. This approach to modelling health outcomes is similar to that described in several other COVID-19 models (Davies et al., 2020b; Keeling et al., 2020).

We use data on the age structure of the Dhaka population (Bangladesh Bureau of Statistics, 2011) (supplementary Table S1) to inform parameters describing the risks of SARS-CoV-2 infections. Previously estimated age-specific risk of death due to COVID-19, of hospitalisation for symptomatic cases (Davies et al., 2020b), and of developing symptoms (Davies et al., 2020a) (supplementary Table S2), were combined with the age distribution in Dhaka District, to estimate the overall proportions of 1) asymptomatic infections, *f*^*a*^ = 0.701, 2) symptomatic infections that lead to death, *f*^*D*^ = 0.009, and 3) symptomatic infections requiring hospitalisation, *f*^*H*^ = 0.073. Of those hospitalised we assume that the proportion requiring critical care in an ICU is *f*^*ICU*^ = 0.31 (World Health Organisation (WHO), 2020a). We assumed no age-structure in contacts within our model.

The model was initially developed as an interactive epidemiological teaching tool (http://boydorr.gla.ac.uk/BGD_Covid-19/CEEDS/), to allow policymakers to explore the impact of interventions on health outcomes and working days lost. For speed and efficiency, and given the large population (Dhaka District population in 2020 is around 13.8 million (Bangladesh Bureau of Statistics, 2011; United Nations, Department of Economic and Social Affairs, 2019b)), the decision was made to make the model deterministic rather than stochastic. We minimise computational complexity by modelling transmission at the population level rather than at the level of the individual or household. However, to more accurately model household quarantining, we further subdivide the six disease states (Fig. 1) to track within-household transmission and account for household-level susceptible depletion (Supplement A).

The ODEs comprising the model are provided in Supplement A and the model parameter descriptions, values and sources, are listed in supplementary Table S4. Analyses were implemented in R (R Core Team, 2019), with the ODEs numerically integrated using the package deSolve (Soetaert et al., 2010). Code can be accessed from our Github repository (https://github.com/boydorr/BGD_Covid-19/BGD_NPI_model).

### Interventions

We implemented three main NPIs – lockdown, household quarantining delivered through Community Support Teams (CSTs), and mask wearing – that were considered to be feasible by policymakers in Bangladesh. For each intervention we considered timing of implementation, scaling up and levels of compliance. In real-time we explored these interventions with policymakers including the lockdown duration, however here we model lockdown as implemented (26^th^ March to 1^st^ June 2020) and consider additional lockdown extensions, which, despite their impracticality, allow comparison to more feasible scenarios.

We define a lockdown as a scenario where all except essential workplaces are closed, including educational facilities, and people are asked to stay home where possible and practice social distancing. For compliant individuals and those that are not essential workers, this intervention is assumed to reduce contacts, and therefore transmission, outside of the household by the proportion *ε*^*ld*^, while leaving within-household transmission unchanged. We achieve this by breaking down the transmission rates for the three infectious states into within- and between-household components using estimates of the SARS-CoV-2 household secondary attack rate, *σ* = 0.166 (Madewell et al., 2020), and the mean household size, *η* = 4 (Bangladesh Bureau of Statistics, 2016). We considered a scale-up period for the lockdown, during which compliance increased linearly from zero to a maximum. Following scale-up, we assume compliance starts to decline exponentially towards a minimum. Full details of implementation are given in Supplement A.

Under the household quarantine intervention, when a symptomatic individual occurs, that individual’s entire household is required to quarantine for 14 days. Those who develop symptoms can self-report to a national COVID-19 hotline or are identified by word-of-mouth, triggering a visit by CST, a volunteer workforce of community-based support workers trained by BRAC/FAO. The CST confirm that symptoms are syndromically consistent with COVID-19, provide information on how to limit spread, facilitate access to healthcare, and offer support to aid quarantine compliance, with additional follow-up during the quarantine period.

Modelling household quarantine requires us to both identify both those individuals that trigger quarantining (the first symptomatic individuals within households), and other individuals in their households who also undergo quarantine. We subdivide the disease states to identify those not currently in an infected household, those in a non-quarantined infected household, and those in a quarantined household. When a susceptible individual in an uninfected household becomes latently infected (through between-household transmission), *η* -1 other individuals also move from uninfected household categories into categories that identify them as being in a non-quarantined infected household, making them vulnerable to both within-household and between-household transmission. We track the group of individuals who were the first infected within a household separately to those who subsequently became infected, allowing us to observe when household index cases become symptomatic. A proportion of these symptomatic index cases comply with quarantine, taking *η* -1 individuals from non-quarantined infected households with them. We also trigger quarantines when within-household cases resulting from index asymptomatic cases become symptomatic. The equations generating these dynamics are described in Supplement A. Infectious individuals within quarantined households are assumed to not cause any between-household transmission. As with lockdown, household quarantining with CST support has defined start and end times, and the proportion of households that comply increases linearly through a scale-up period, after which compliance remains at a constant maximum (see Supplement A).

Finally, we considered compulsory mask-wearing outside of the household. Within the model, masks are assumed to block a proportion, *ε*^*m*^, of between-household transmission from compliant individuals, while also blocking a proportion, *ε*^*m*^*ρ*^*m*^, of transmission to compliant individuals, where 0≤*ρ*^*m*^ ≤1, i.e. masks protect others from the wearer, and the wearer from others, to different degrees, with protection provided to the wearer never greater than that provided to others (Howard et al., 2020; Stutt et al., 2020). The full description of mask wearing effects, in relation to timing and compliance is provided in the Supplement A.

We model working days lost due to both illness and interventions. Based on the 2011 census (Bangladesh Bureau of Statistics, 2011), we assume 52% of the population is formally employed, working five days a week. We model symptomatic and recuperating individuals to be off-sick, and assume that deaths result in loss of all subsequent working days through 2020. During lockdown, those workers that are both compliant and not essential workers lose their working days. Those in quarantined households do not work, and, since household quarantine is based on symptoms, we assume that, in addition to those quarantined due to COVID-19, a proportion of households affected by non-COVID-19 influenza-like illnesses also undergo quarantine. Based on the 2011 census, 23% of the population is unemployed, but works within the household, e.g. with caring responsibilities (Bangladesh Bureau of Statistics, 2011). When these individuals are hospitalised or die, we assume another (possibly formally employed) household member replaces them, leading to further loss of working days. Finally we assume that, in response to each death, a number of grieving individuals (taken to be *η* -1) do not work for a week. Details of the working days lost calculation are provided in equations S.16-17.

### Scenario comparison

With input from policy-makers, we explored 15 combinatorial scenarios for 2020, including a baseline with no interventions and unmitigated SARS-CoV-2 spread.

The first of four lockdown-only scenarios aimed to replicate the lockdown as implemented, from 26^th^ March until 1^st^ June 2020. The other three lockdown-only scenarios involved extending the lockdown by 1, 2 and 3 months. In all four scenarios we assume scale-up occurs over one week, with peak compliance of 80%, declining towards 30% (supplementary Fig. S1D); considered to be qualitatively similar to practice in Dhaka. The impact of lockdown on between-household transmission of compliant individuals *ε*^*ld*^ was estimated during model calibration (see below).

We also examined scenarios where lockdown, as implemented in Bangladesh, was followed by either CST interventions, compulsory mask-wearing or both, beginning a week before lockdown ended and continuing through 2020 with a 7-day scale-up period and 80% peak compliance with each intervention. The ability of masks both to protect others from transmission from the wearer and to protect the wearer from transmission from others depends on several variables, including the material used, the construction (e.g. layers of material), and the quality of fit (Aydin et al., 2020; Davies et al., 2013; Howard et al., 2020). Therefore, for mask-wearing we consider scenarios where there is low, medium, and high protection of others from the wearer (*ε*^*m*^ ={0.2, 0.5,0.8}), and where the protection provided to the wearer is zero, half that provided to others, or equal to that provided to others (*ρ*^*m*^ ={0,0.5,1}), giving nine mask-wearing scenarios.

For the 2020 time horizon we compared the total: 1) hospitalisations, 2) deaths, 3) percentage of patient days exceeding hospital capacity (the sum of patients in excess of hospital beds each day, divided by the summed total patients over all days), 4) working days lost, 5) cost of implementing interventions and of healthcare for COVID-19 patients, 6) cost per death averted (relative to no intervention), and 7) the percentage return (in terms of healthcare savings) on investment (%ROI) in interventions. For estimating total cost we include healthcare provision for hospitalised COVID-10 patients, media campaigns, training and deployment of CSTs, and mask distribution, as detailed in Table 1. For each scenario, we used the total cost to estimate the cost per death averted. This was calculated by subtracting the cost of the baseline scenario from that of the focal scenario, then dividing by the reduction in deaths from the baseline. The %ROI for each scenario was calculated by subtracting the total cost of each scenario from the baseline, and dividing by the intervention implementation costs (i.e. the sum of all costs other than hospital care).

**Table 1:**
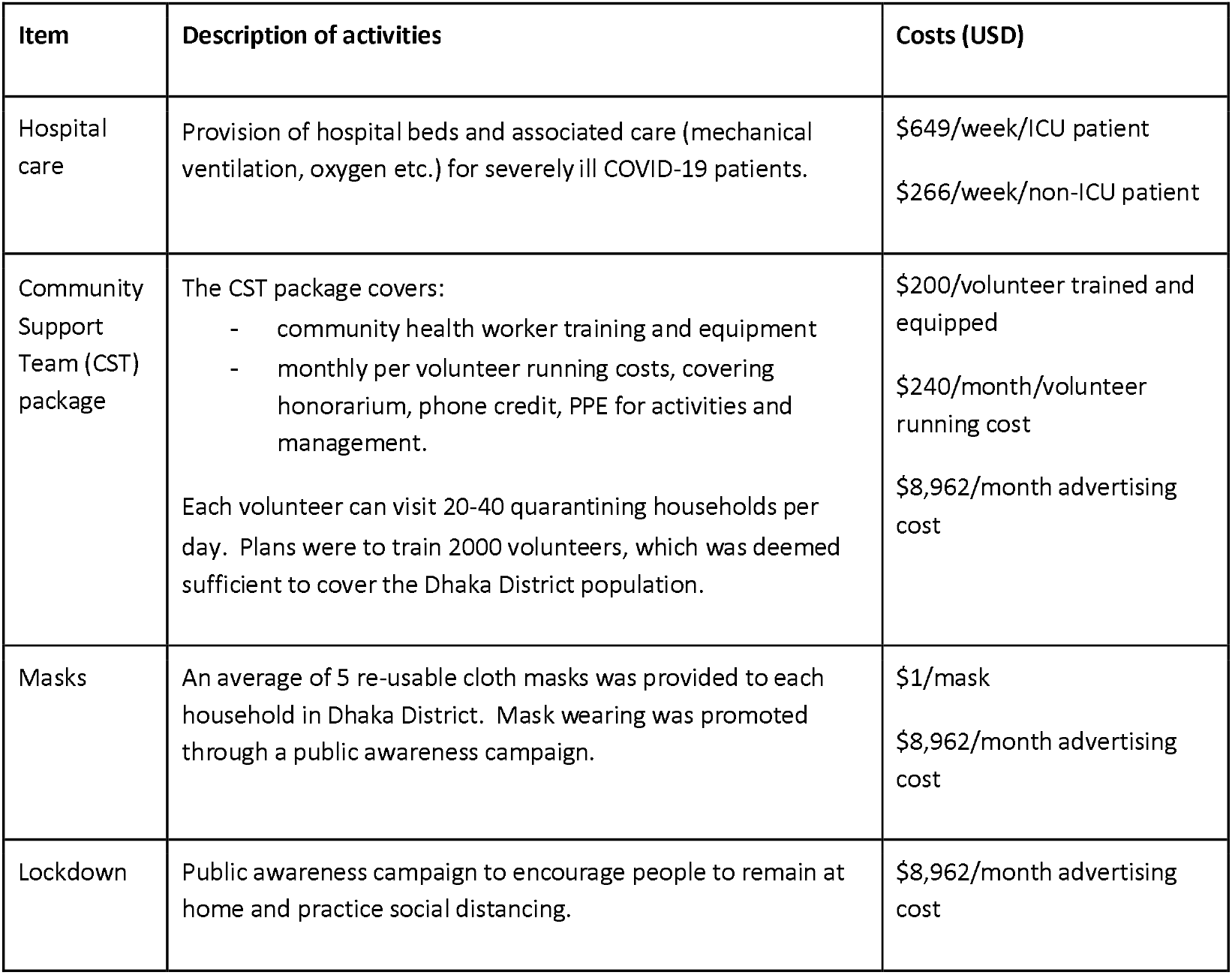
Costs associated with healthcare and interventions, derived from ongoing practices in Bangladesh.

| Item | Description of activities | Costs (USD) |
| --- | --- | --- |
| Hospital care | Provision of hospital beds and associated care (mechanical ventilation, oxygen etc.) for severely ill COVID-19 patients. | \$649/week/ICU patient<br>\$266/week/non-ICU patient |
| Community Support Team (CST) package | The CST package covers:<br>- community health worker training and equipment<br>- monthly per volunteer running costs, covering honorarium, phone credit, PPE for activities and management.<br>Each volunteer can visit 20-40 quarantining households per day. Plans were to train 2000 volunteers, which was deemed sufficient to cover the Dhaka District population. | \$200/volunteer trained and equipped<br>\$240/month/volunteer running cost<br>\$8,962/month advertising cost |
| Masks | An average of 5 re-usable cloth masks was provided to each household in Dhaka District. Mask wearing was promoted through a public awareness campaign. | \$1/mask<br>\$8,962/month advertising cost |
| Lockdown | Public awareness campaign to encourage people to remain at home and practice social distancing. | \$8,962/month advertising cost |

Given the heavy societal and economic costs of lockdowns, they are typically viewed as temporary measures to buy time for implementation of other less restrictive measures (e.g. mask wearing) or more targeted measures (e.g. household quarantines and contact tracing). To explore the impact of preparedness prior to lockdown ending, we examined the sensitivity of model outcomes (hospitalisations, deaths, working days lost, and exceeded hospital capacity) to the scale-up period and start date of post-lockdown interventions. Modelling lockdown as implemented in Bangladesh, we looked at scenarios where lockdown was followed by household quarantining only, mask-wearing only (assuming mask protectiveness parameters *ε*^*m*^ = 0.5 and *ρ*^*m*^ = 0.5), or both. We varied the start date of the post-lockdown intervention(s) from 30 days pre-to 30 days post-lockdown ending, while keeping the scale-up period constant at 7 days, and vice versa varying the scale-up period (zero to 30 days), while keeping the start constant (7 days prior to the lockdown end).

### Model calibration

We calibrated two parameters, *R*_0_ and *ε*^*ld*^, the proportion by which lockdown reduces between-household transmission from compliant people, against daily COVID-19 death data in Bangladesh (European Centre for Disease Prevention and Control, 2020). We chose to calibrate against reported deaths rather than cases, since death data are less affected by testing and reporting capacity. To obtain a time-series of deaths in Dhaka District, we extracted the proportion of district-resolved case totals for Dhaka District from Bangladesh’s COVID-19 dashboard (“Coronavirus COVID-19 Dashboard, 2020,” 2020) and assumed this proportion remained constant. The first three cases in Bangladesh were confirmed on 8^th^ March 2020, but it is thought likely that infection began circulating undetected prior to this, with genomic data indicating an introduction in mid-February and at least eight introduced cases prior to the ban on international travel (Cowley et al., 2021). We therefore initialise the model with eight infectious cases on 15th February 2020. The value of *R*_0_ was optimised to minimise the absolute difference between modelled and reported cumulative deaths on the date lockdown started (26th March). Following this optimisation of *R*_*0*_, the value of *ε*^*ld*^ was similarly estimated by minimizing the difference between modelled deaths and data on the date lockdown ended.

We recalibrated the model to data (Krispin and Byrnes, 2021) from the rapid resurgence beginning in March 2021 to learn about the likely *R*_0_ at that time, when B.1.351 increased in frequency to become the dominant virus variant (Saha et al., 2021). In March 2021, levels of mask-wearing and compliance with household quarantine were low, and there was no lockdown in place. As cases increased, a loose lockdown was introduced on 5^th^ April, with stronger restrictions applied on 14^th^ April and maintained until end May, with some relaxing of restrictions over this period. We modelled this with a lockdown from 5^th^ April to 1^st^ June, with a 9 day scale-up period, maximum compliance of 80%, and the same rate of declining compliance as in 2020 (supplementary Fig.S1C). We also assume that from 5^th^ April to 1^st^ June there is an increase in both mask-wearing (with *ε*^*m*^ = 0.5 and *ρ*^*m*^ = 0.5) and household quarantine, with a scale-up period of 9 days to reach a compliance of 30%. Given considerable uncertainty around the number of infectious and immune individuals in the population, we calibrated the model under a range of initialisation scenarios in terms of immunity levels (0%, 25% and 50%) and circulating cases on 1^st^ March. We assumed that reported daily new cases represented around 10% of true cases (supplementary Fig. S1C), and that prevalent infectious individuals were about eight times incident daily cases given the infectious period duration. Dividing by two, assuming that these individuals are on average halfway through their infectious period, we get an effective number of 11,451 infectious individuals for initialization and an equivalent number of incubating individuals, but we also considered numbers 50% higher and lower than this. We took two approaches to optimisation of *R*_0_, minimizing the difference: 1) between modelled and reported cumulative deaths over the period from 1^st^ March-5^th^ April 2021, and 2) in the timing of the peaks in reported cases and modelled symptomatic cases (with the constraint that modelled cumulative deaths from 1^st^ March until 20^th^ May had to be at least as high as reported deaths over the same period).

### Sensitivity Analyses

There is considerable uncertainty in the parameters governing SARS-CoV-2 transmission and health impacts. We therefore undertook one-way sensitivity analyses across parameter value ranges for *R*_0_, the duration of disease states and health outcome stages, the introduction date, the household secondary attack rate, the ratios of asymptomatic to symptomatic transmission, and of pre-versus symptomatic transmission identified as plausible from the literature (supplementary Table S4). The impact of changes in each of these parameters on the main model outputs was assessed individually, while keeping other parameter values fixed. We ran this analysis over scenarios of: 1) no interventions; 2) lockdown as implemented; 3) lockdown plus household quarantining; and 4) lockdown plus compulsory mask wearing (with *ε*^*m*^ = 0.5 and *ρ*^*m*^ = 0.5).

## Results

### Model calibration

We estimated that *R*_0_ = 3.57 for transmission of SARS-CoV-2 in Dhaka in 2020, and that the lockdown reduced between-household transmission (*ε*^*ld*^) by 97% for compliant individuals. The match between modelled deaths and data during the early stages of the epidemic is illustrated in supplementary Fig. S1A and B). Outputs from the calibrated model until the lockdown end indicate that less than 10% of cases were recorded during this period (supplementary Fig. S1C). We further explored how testing capacity might impact case detection within our interactive app, which similarly indicated that the low, but increasing, testing capacity in Dhaka would fail to detect a high proportion of cases given the high proportion of asymptomatic cases and distribution of tests to individuals with other influenza-like illnesses.

Our estimates of *R*_0_ in 2021 varied based on the initialisation scenario, with values increasing with lower initial circulating cases and higher levels of immunity in the population (supplementary Table S5). The approach used to optimise *R*_0_ also impacted the estimates, with optimisation based on matching to deaths in the early stages of the resurgence leading to lower values (range of 2.19-6.06) compared to peak matching (range of 3.90-8.20). Our best guess initialisation scenario (11,451 initial infections and 25% prior immunity), led to an *R*_0_ estimate of 3.32 when matching to deaths and 5.30 when matching to peak timing, and corresponding death detection estimates of 32-75% (matching to deaths), or 8-22% (peak matching). The peak in symptomatic cases obtained by matching to deaths was always slightly later than the peak in reported cases, though never by more than four days. When calibrated to the trajectory of deaths, the model predicted that while NPIs controlled the resurgence, another peak will occur in the coming months following their relaxation (supplementary Fig. S2C). In contrast, calibration by matching peak timing leads to the prediction that the resurgence sufficiently spread through the remaining susceptible population to prevent further waves (supplementary Fig. S2D).

### Impacts of scenarios on health outcomes and working days lost

In the absence of interventions we predicted that COVID-19 patients would peak at 46,309 in late May (Fig. 2A), greatly exceeding hospital bed capacity (estimated to be 10,947 in Dhaka District (World Health Organisation (WHO), 2020b); supplementary Table S4). Under this scenario hospital beds would have been unavailable for at least 55% of patient days (Fig. 3D) without accounting for non-COVID-19 bed needs. Assuming unmitigated transmission, the epidemic would have likely led to around 13.3 million cases (i.e. most of the Dhaka District population; supplementary Table S4) and 35,849 deaths (Figs. 3A, S2A & S3A). Although high relative to some of the mitigated scenarios, these deaths represent 0.26% of the population, and would be expected to lead to a loss of <1% of total working days (Fig. 3C).

**Figure 2:**
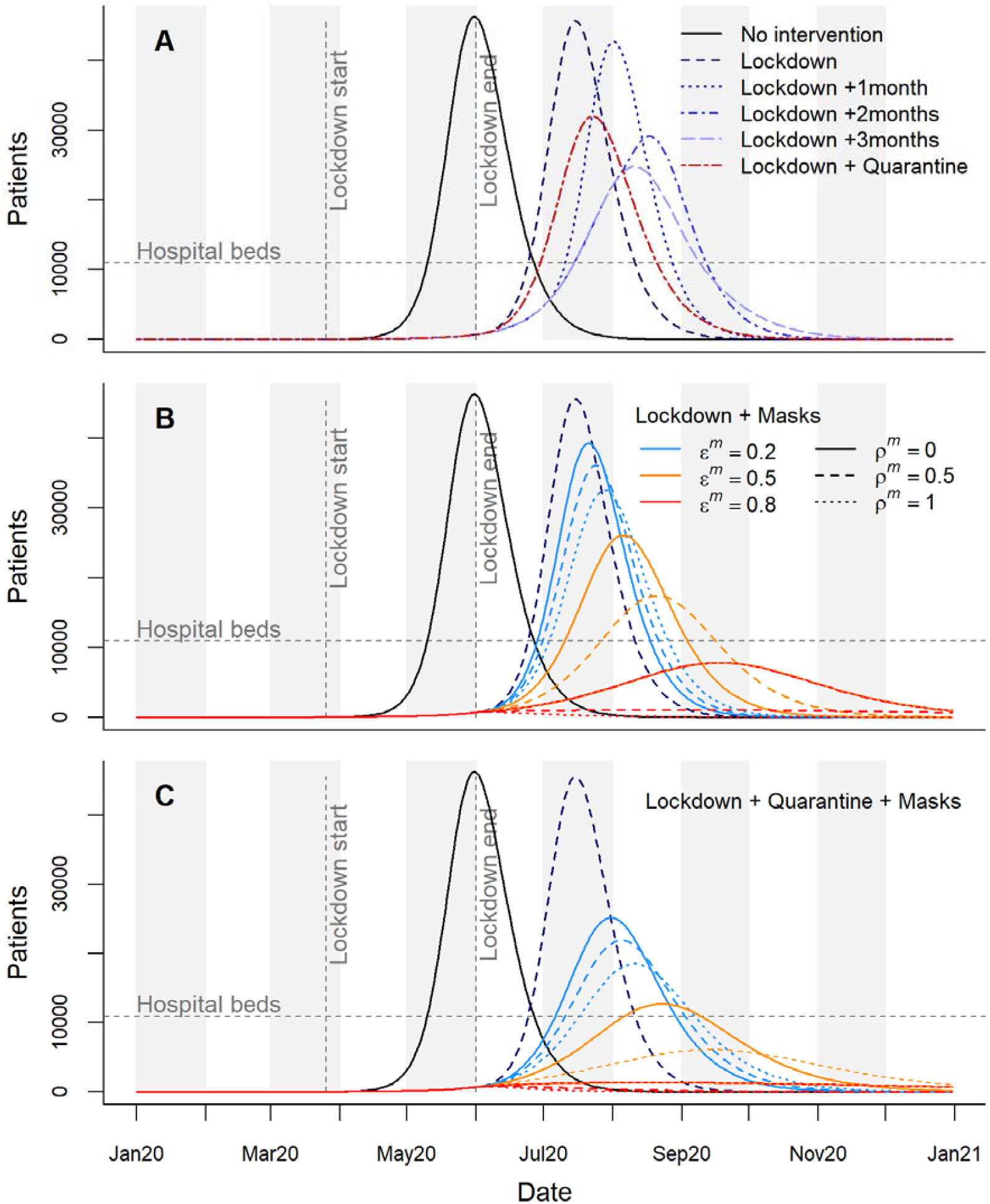
Time series of hospital patients for intervention scenarios in 2020. Horizontal dashed lines indicate hospital bed capacity in Dhaka District. Vertical lines indicate the start and end points of lockdown as implemented in Bangladesh. A) Hospital patients in the absence of interventions, for the implemented lockdown plus extensions of up to 3 months, and for lockdown followed by household quarantine with community support teams. B) Lockdown as implemented followed by compulsory mask wearing, considering nine mask effectiveness scenarios; *ε*^*m*^ describes the proportion reduction in outward emissions by mask wearers, while *ρ*^*m*^*ε*^*m*^ describes the proportion protection to mask wearers from others’ emissions. C) Combined impacts of the lockdown, household quarantine, and masks of different effectiveness.

**Figure 3:**
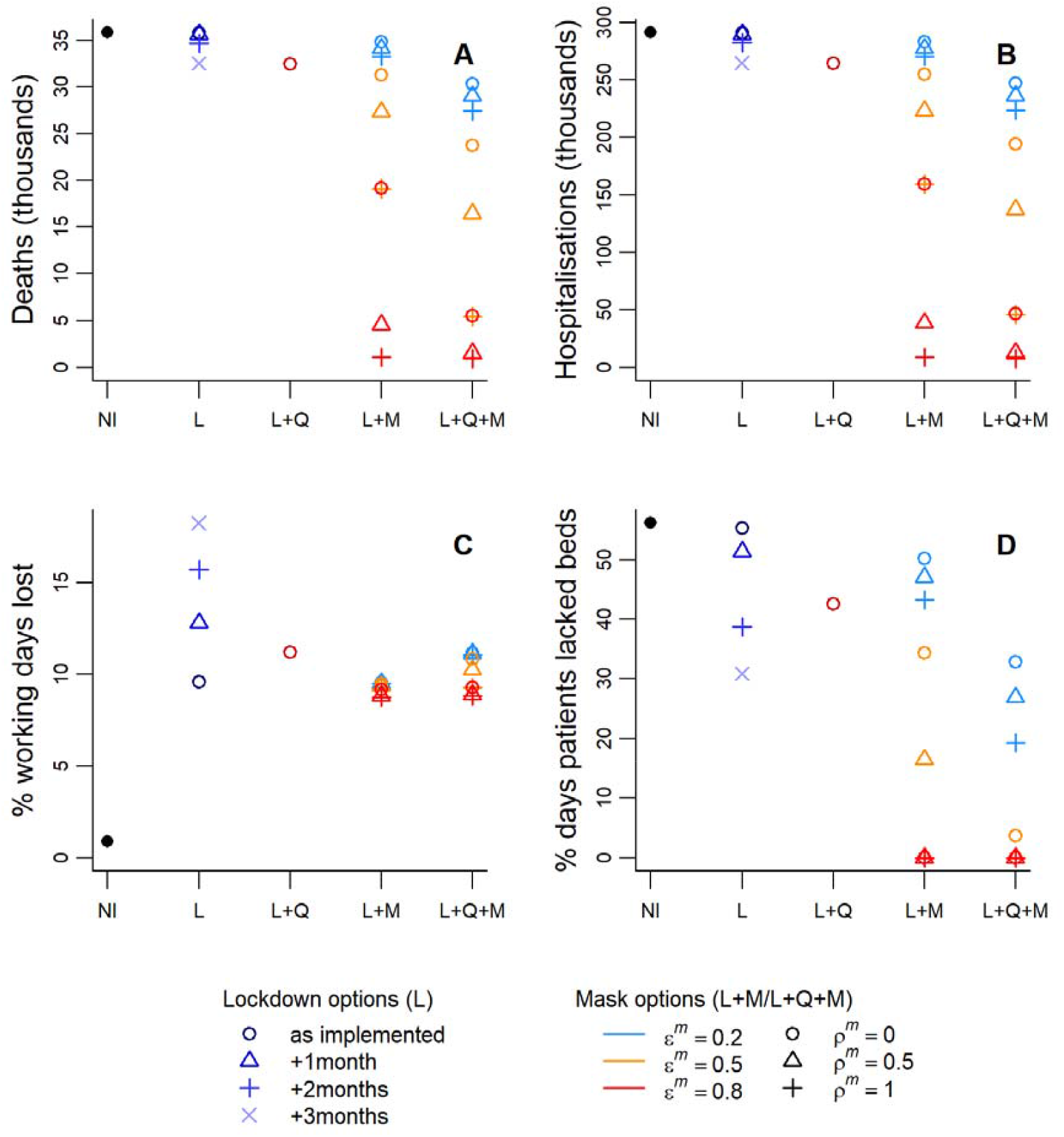
Summaries of health outcomes and working days lost for a range of interventions during 2020. NI=no intervention, L=lockdown, Q=household quarantine with community support teams, M=compulsory mask wearing. *ε*^*m*^ describes the proportional reduction in outward emissions by mask wearers, while *ρ*^*m*^*ε*^*m*^ describes the proportion protection provided to the mask wearer from others’ emissions.

We forecast that the lockdown as implemented in Bangladesh would delay the epidemic (Figs 2A and S2A), with minor impacts on deaths and hospitalised cases (Fig. 3), while increasing working days lost to 9.6%. Extensions were predicted to delay and widen the peak (Figs 2A and S2A, extensions shown for up to three months, as further extensions had little impact on the epidemic peak or total outcomes). Even with lockdown extensions, hospitalisations were still expected to outstrip capacity (Fig.2A), with only modest reductions in deaths (Fig.3). Working days lost were predicted to increase by 2.5-3.5% with each extra month of lockdown (Fig.3C).

We found that introducing household quarantining following lockdown led to the epidemic peak being later, lower and wider than with lockdown alone (Figs 2A and S2A). Hospital capacity was still exceeded, with deaths and hospitalisations reduced to levels similar to those achieved by a three-month extension to lockdown. The additional working days lost by introducing and maintaining household quarantines throughout 2020 were fewer than those lost by a one-month lockdown extension (Fig. 3C).

The impact of introducing compulsory mask-wearing following lockdown varied based on the effectiveness of the masks used (Figs 2B and 3). When *ε*^*m*^ = 0.2, i.e. low filtration efficiency, only relatively small reductions to the epidemic peak were predicted, with the decline increasing with mask effectiveness in terms of PPE (indicated by increasing *ρ*^*m*^ ; Figs 2B and S2B). Masks with high filtration efficiency (*ε*^*m*^ = 0.8), or medium filtration efficiency combined with high PPE efficiency (*ε*^*m*^ = 0.5, *ρ*^*m*^ =1) flattened the peak sufficiently to keep patient numbers below hospital capacity (Figs 2B and 3D). Effective masks also led to substantial drops in deaths and hospitalisations, while slightly reducing working days lost relative to the lockdown-only scenario (Fig. 3).

Combining mask-wearing with household quarantine led to greater predicted reductions in the epidemic peak and in both deaths and hospitalisations than either intervention alone (Figs 2C and 3A-B). Percentages of working days lost increase as mask quality decreases, ranging from 8.8-11.2%. A combined strategy of masks and quarantine following lockdown was strategized for implementation in Dhaka District. In the end, mask provision was not directly funded despite recommendations, but mask-wearing was promoted; by late 2020 mask use had largely declined to normal levels. The data-based estimates of cumulative deaths in Dhaka District take a path similar to those modelled by combining quarantining with either masks of high filtration effectiveness (*ε*^*m*^ = 0.8) and low to medium PPE abilities (*ρ*^*m*^ of 0 or 0.5), or with masks of medium filtration effectiveness (*ε*^*m*^ = 0.5) and *ρ*^*m*^ =1 (supplementary Fig. S4C).

### Scenario costs

Total costs of the scenarios explored ranged from $20.2.0-111.4 million (Fig. 4A). Despite requiring no direct intervention costs, the baseline scenario is among the most expensive due to the healthcare costs incurred by hospitalisations. Lockdown as implemented was predicted to be similar in cost to the baseline (unmitigated transmission) with modest declines for lockdown extensions. Incorporating household quarantine led to costs similar to extending lockdown for 2-3 months. Combining lockdown with low effectiveness masks led to the highest costs due to the expense of mask distribution, offset by only small reductions in healthcare costs. Masks of high effectiveness, however, led to substantial cost reductions, which generally increased when masks and household quarantine were modelled together.

**Figure 4:**
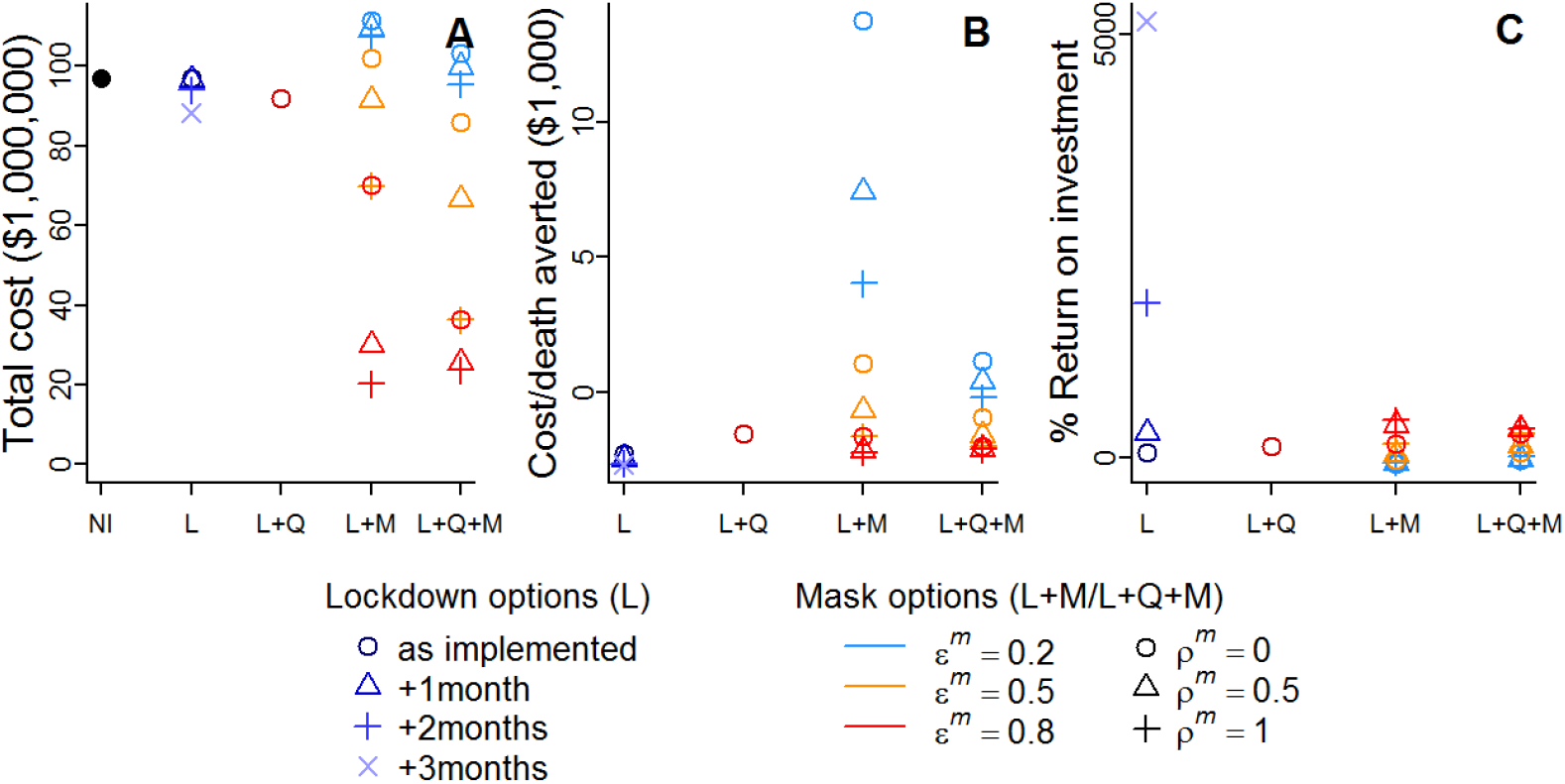
Costs of intervention scenarios. NI=no intervention, L=lockdown, Q=household quarantine with community support teams, M=compulsory mask wearing. *ε*^*m*^ describes the proportional reduction in outward emissions by mask wearers, while *ρ*^*m*^*ε*^*m*^ describes the proportion protection provided to the mask wearer themselves from others’ emissions. A) Total cost of each intervention scenario to the government based on costings in Table 1. B) Cost of each death averted relative to the baseline (NI) scenario. C) Percentage return on investment in interventions in terms of healthcare savings.

Since most scenarios had a lower total cost than the baseline, the cost per death averted is negative except in some of the lower effectiveness mask scenarios (Fig. 4B). Extended lockdowns, and lockdowns combined with household quarantine and/or high effectiveness masks all provide similar savings per death averted in the $1,500-$2,700 range.

The %ROI was generally positive, again with some exceptions where low effectiveness masks were used (Fig. 4C). By far the highest %ROIs were given by extending lockdown by 2-3 months. This is because, despite relatively small reductions in hospitalisations (Fig. 3B), the only costs to lockdown extension are in advertising, and these costs are small relative to those of providing masks to every household or maintaining CST (Table 1). Lockdown plus quarantining gave a %ROI of 127%. The %ROI of all scenarios involving masks increased with the effectiveness of the masks in blocking transmission. Interventions involving high effectiveness masks all had a higher %ROI than the lockdown plus quarantine scenario.

### Intervention timing and scale-up

Starting either household quarantining or compulsory mask-wearing prior to the end of lockdown had little projected impact on health outcomes or the percentage of working days lost (Fig. 5A-D). When masks and quarantining are combined, however, total deaths and hospitalisations are seen to decline in response to moving their start dates further before the end of lockdown. This difference occurs because the combined early start date of the two interventions pushes the epidemic peak later (into 2021) than when considering either individually. Continuing interventions and calculating outcomes over both 2020 and 2021, removes this apparent improvement in outcomes with start dates prior to lockdown’s end (supplementary Fig. S5A-D). As the start date of quarantining and mask-wearing is delayed beyond the lockdown end date, their health benefits decline, with all outcomes approaching those of the lockdown-only scenario. Similar consequences to delaying intervention start dates result from lengthening the scale-up period (Fig. 5E-H).

**Figure 5:**
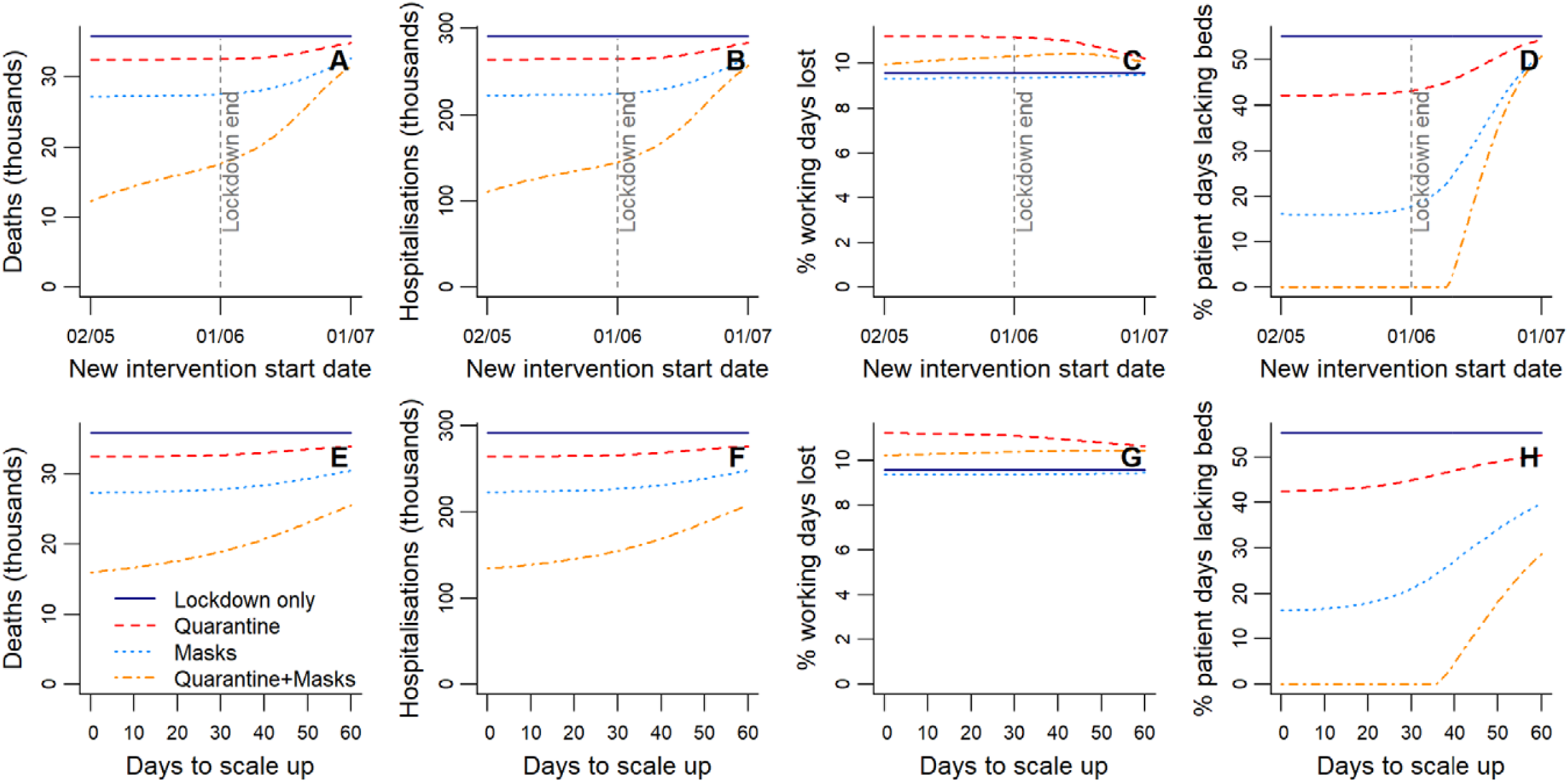
Sensitivity to the start date and scale-up period of quarantine or mask wearing following lockdown. A-D) Changes in health outcomes and working days lost over 2020 when the start date of interventions (household quarantine, masks, or quarantine and masks) following lockdown is adjusted relative to the lockdown end date. The days taken to scale up interventions to their full effectiveness is held constant at seven days. E-H) Changes in the same outcomes when the time in days taken to scale post-lockdown interventions is varied. The start date of the post-lockdown interventions is held constant at seven days prior to the lockdown end date.

### Sensitivity Analyses

*R*_0_ was among the top three most influential parameters for all five outcome measures reported under all four baseline scenarios (Figs. S5-8). The duration of symptoms was among the top three most influential parameters in determining working days lost (since symptomatic people are unable to work) over all baselines, and, predictably, the mean lengths of stay in general hospital and ICU beds were highly influential for the percentage of patient days that lacked beds. For all baselines, other than that with no interventions, the introduction date of the disease ranked in the top three most important parameters for all outcomes except the percentage of patient days lacking beds. Some parameters only gained importance under specific baseline scenarios (for example, the level of asymptomatic transmission becomes important under symptoms-based household quarantine); for more details see Supplement D.

## Discussion

We modelled interventions for controlling COVID-19 transmission in Dhaka District, Bangladesh, comparing them based on their ability to prevent both deaths and overwhelming of the health system, as well as on their societal and health provider costs. We found that under expected compliance, lockdowns alone, regardless of duration, were both costly and unable to keep cases below hospital capacity, while preventing only a small proportion of deaths, confirming that they are not an appropriate long-term measure in this context (Amewu et al., 2020; Andam et al., 2020; Rahman et al., 2020). Additional quarantining of households with symptomatic individuals similarly was not predicted to prevent hospitalisations exceeding capacity. Modelling compulsory mask-wearing after the lockdown produced outcomes that varied widely depending on the effectiveness of masks in blocking transmission. Low-quality masks had limited impacts and were not cost-effective, whereas high-quality masks substantially reduced deaths, protected the healthcare system and were cost-effective. Combining mask-wearing and household quarantine led to further reductions in deaths and excess hospitalisations, and was cost effective when masks were of high quality. Simulations also suggest that early introductions of post-lockdown measures would have had negligible additional impact over the full course of the epidemic (though they may improve outcomes in the short-term), but gaps between lockdown ending and post-lockdown interventions (or long periods of scale-up) quickly dilute their impacts.

The inability of symptomatic household quarantining alone to prevent hospitalisations exceeding capacity is unsurprising. Even in HICs with greater healthcare capacity, lower *R*_0_ estimates, and a lower proportion of asymptomatic transmission, other modelling studies have suggested that symptoms-based household quarantine would still overwhelm health systems (Aleta et al., 2020; Ferguson et al., 2020). Additional tracing and quarantine of non-household contacts of symptomatic individuals can improve the effectiveness of quarantine measures (Aleta et al., 2020). However capacity for extensive contact tracing is likely to be limited in high-density resource-constrained settings like Dhaka. The finding that combining symptomatic household quarantining with mask mandates (using mid-to high-quality masks) leads to effective control was crucial at the time, since both these NPIs were considered feasible. Given that household quarantining is triggered by a symptomatic case, and we estimate that most cases in Bangladesh are asymptomatic, a large proportion of infectious households are still likely missed by this measure, which will also have only limited impact on pre-symptomatic transmission. Since masks reduce transmission for all infections, irrespective of symptoms, masks work synergistically with household quarantine. While we consider scenarios where masks block only 20% of transmission, we note that this is a worst case scenario; experimental work suggests that our mid-to high-quality mask scenarios (blocking 50-80% of transmission) are more likely (Aydin et al., 2020; Davies et al., 2013). Observational (Hong et al., 2020; Wang et al., 2020) and other modelling (Stutt et al., 2020) studies provide further evidence for the effectiveness of masks in blocking transmission.

In reality, achieving the modelled effectiveness of household quarantining and mask-wearing depends on high compliance (we assumed 80% compliance in the main results presented here). A survey in Israel suggested that quarantining compliance is likely to be highly dependent on compensation for loss of work (Bodas and Peleg, 2020). However, such compensation may be unachievable in many LMICs. Reducing insecurity experienced by households under quarantine in other ways, for example by food provisioning and healthcare access, may help mitigate income loss and boost compliance amongst poorer households. In Bangladesh, CSTs are already playing this role in supporting quarantining households, and in LMICs more broadly, community health workers are likely to prove invaluable in using the trust they have established to encourage compliance with NPIs (Ballard et al., 2020).

Most of the NPIs we explored were very cost-effective, with savings per death averted and a positive %ROI as a consequence of the high contribution of healthcare to overall costs. Note that these returns occur despite the relatively young population in Bangladesh, which leads to a low proportion of hospitalized cases (0.022). Furthermore, the societal costs (in terms of working days lost) of the post-lockdown NPIs considered were small when compared with the initial lockdown costs and potential extensions. These findings are in line with other work showing that the costs of unmitigated transmission exceed those of implementing NPIs (Thunström et al., 2020; Torres-Rueda et al., 2020). The costs we explore here are based on crude assumptions for mask purchase and costs of training and rollout of CSTs in Dhaka where there is already a large community health workforce that can be mobilized. However, we do not include food packages or additional support for vulnerable communities that may increase NPI effectiveness but also costs.

Throughout the development of our model and interactive app, we incorporated suggestions from policymakers on questions that most urgently needed answers, and on NPIs under consideration and thought feasible to implement. This co-development allowed the investigation of scenarios appropriate to the local context that addressed pressing policy concerns (McBryde et al., 2020). The app, which allows a user-friendly exploration and visualisation of how scenarios impact health outcomes and costs, proved to be an effective tool to support discussions with policymakers, as the timing and combination of interventions, along with uncertainties in parameters, including compliance, could be explored on the fly. This proved to be crucial in understanding the economic and health trade-offs involved, as well as helping to demystify the model itself and the ensuing epidemic.

Our study has a number of limitations. First, to ensure our model could run sufficiently quickly within the interactive app, stochasticity and individual variation in transmission was not incorporated. We expect only a minimal impact of these simplifications on disease trajectories and key results due to the large population size considered and rapid epidemic growth observed. However, they do limit the usefulness of the model for exploring elimination scenarios, since stochasticity and superspreading, inherent to SARS-CoV-2 transmission (Adam et al., 2020), become more influential under low levels of infection (Vespignani et al., 2020). Imported infections, which may similarly become important near elimination, were also not considered. Age-structured models have previously been proposed for studying COVID-19 (Davies et al., 2020b; Ferguson et al., 2020; Keeling et al., 2020). We did not include age-structured transmission, which though not entirely realistic, may be reasonable given the high degree of intergenerational mixing in Bangladesh (United Nations, Department of Economic and Social Affairs, 2019a) and some other LMICs (Hodgins and Saad, 2020). We also do not consider exacerbated mortality when hospital capacity is exceeded, potentially underestimating mortality under some scenarios. Parameters used for case fatality, and proportions of symptomatic and hospitalised infections, were based on age-dependent estimates from HICs (Davies et al., 2020b, 2020a) and the age-distribution in Dhaka District. However, these HIC-derived parameters may be less accurate when applied in this setting, as the incidence among age classes of underlying conditions that increase COVID-19 risk likely differs in LMICs (Clark et al., 2020). We also note that our estimates of working days lost make the assumption that employed people cannot switch to working from home, possibly leading to overestimation.

More generally there remains considerable uncertainty around many of the model parameters. Our sensitivity analyses indicated that *R*_0_ was, predictably, very influential in determining health outcomes, but is sensitive to the introduction date and number of imported cases which are uncertain. In addition, prior to the lockdown in 2020, only 5 deaths due to COVID-19 had been recorded in Bangladesh, and given the inherent stochasticity in these events, tuning *R*_0_ to data in this time window is unlikely to be very accurate. Finally, prior to the 2020 lockdown, some control measures, such as cancellations of large gatherings, had already been taken(Anwar et al., 2020), potentially lowering our *R*_0_ estimate. Our estimates of *R*_0_ in 2021 similarly assumed no interventions in the run-up to the 5^th^ April lockdown, but low levels of mask-wearing and quarantine may have led to underestimation. The 2020 *R*_0_ estimate for Dhaka lies within the 90% confidence interval estimated from a meta-analysis of pre-March 2020 estimates (Davies et al., 2020b). Many of our 2021 estimates exceed this confidence interval, but none are in excess of the confidence interval reported by Sanche et al. (2020). A possibly higher *R*_0_ value for 2021 is also not unexpected, given the dominant variant at the time (B.1.351) was known to be more transmissible than variants prevalent in 2020. The effect of lockdown on between-household transmission rates was, like *R*_0_, tuned to reported death data, and proved to be higher than was anticipated in this high density population; reducing transmission by 97%, suggesting that lockdown was very effective as a short-term control measure. This high impact on compliant individuals might have been compensating for our conservative assumption of no reduction in transmission from essential workers under lockdown (unless off sick with symptomatic COVID-19), or underestimated lockdown compliance. An overestimated *R*_0_ (or indeed inaccuracies in other parameters) could also lead to overestimated impacts of lockdown.

Our initial model assumed that recovered individuals remain immune through 2020. Although immunity to SARS-CoV-2 is not permanent (Iwasaki, 2021), this assumption appeared reasonable given effects of immunity loss were likely limited over this period. However, with resurging cases in 2021 concomitant with relaxed NPIs and the emergence of the more transmissible B.1.351 variant, the question of immunity became paramount (Saha et al., 2021). The level of immunity in the population at the time of the resurgence was very uncertain, given only limited data from which to infer case detection and laboratory evidence that prior COVID-19 infections elicit less protection to the B1.351 variant (Planas et al., 2021). These uncertainties translated into our estimates of *R*_0_ in 2021, which varied widely (2.19-8.2) under different assumptions, but with a best guess of 3.32-5.30. The uncertainty in *R*_0_ in turn leads to uncertainty in predictions, with some suggesting a possible further wave later in 2021, while others preclude this.

We demonstrated the sensitivity of outcomes to the timing and scale-up of interventions, but human behavioural responses most dramatically impact outcomes. For these reasons, our model was primarily developed as a means to understand the potentially synergistic impact of interventions, rather than to accurately forecast dynamics subject to unpredictable changing human behaviours. We therefore considered compliance to interventions to be a crucial interactive element of our app to build understanding and guidance on policy. Within the app, we also modelled the degree to which the limited (but greatly increased) testing capacity would still under-detect circulating cases, given some degree of cognitive dissonance and the considerable uncertainty in pre- and asymptomatic transmission during the early months of the pandemic. Overall we found the interactive app to be effective for communicating epidemiological modelling outcomes to policymakers together with their caveats, and we recommend the use of such tools that can be tailored to other settings and interventions.

In summary, we found that two NPIs combined, masks and symptoms-based household quarantining, were capable of averting an anticipated public health crisis in Dhaka, while also being good value for money. These measures were to a large extent rolled out in Bangladesh in 2020, and appear to have contributed to limiting transmission, but the ensuing epidemic stretched the health system. In practice, compliance with these interventions and fidelity of their implementation was highly heterogeneous, with measures relaxing over the year as activity returned to levels approaching normalcy. The surge in cases in 2021, apparently driven by the B.1.35 variant, has now largely subsided. However, predictions about the risk of a further wave depend on, as yet, unqualified assumptions and vaccination coverage remains low. NPIs therefore still have an important role to play, particularly mask wearing, which is both very cost-effective and feasible, and vaccine rollout needs to be pursued at pace.

## Data Availability

All data and code can be accessed via out Github repository (https://github.com/boydorr/BGD_Covid-19/BGD_NPI_model). The interactive app is available at http://boydorr.gla.ac.uk/BGD_Covid-19/CEEDS/.

http://boydorr.gla.ac.uk/BGD_Covid-19/CEEDS/

## CRediT authorship contribution statement

**Elaine A Ferguson:** Conceptualization, Data curation, Formal analysis, Investigation, Methodology, Software, Visualization, Writing – original draft, Writing – review & editing. **Eric Brum**: Conceptualization, Funding acquisition, Investigation, Project administration, Writing – review & editing. **Anir Chowdhury:** Investigation. Shayan Chowdhury: Investigation. **Mikolaj Kundegorski:** Data curation. **Ayesha S Mahmud**: Methodology, Writing – review & editing. **Nabila Purno**: Data curation, Investigation, Project administration. **Ayesha Sania:** Funding acquisition, Investigation. **Rachel Steenson**: Data curation, Software, Visualisation. **Motahara Tasneem**: Data curation, Project administration. **Katie Hampson**: Conceptualisation, Funding acquisition, Data curation, Investigation, Methodology, Software, Supervision, Writing – original draft, Writing – review & editing.

