## Supplementary Information for "Modelling to inform the COVID-19 response in Bangladesh"

### Supplement A: Model description

The model as outlined in Fig. 1 is composed of ordinary differential equations (ODEs) describing the changing population in each of the disease and health outcome states. We introduced some sub-division of the six main disease states in order to implement household quarantining, where the quarantined population is prevented from transmitting to the non-quarantined population. The equations defining the model are described in this supplement. Descriptions of all modelled state variables can be found in Table S3, and descriptions of the parameters and their assumed values (with sources) are provided in Table S4.

Within our model, we divide the population that is susceptible to SARS-CoV-2 infection (denoted $S$) into four sub-categories ($S = S^n + S^E + S^I + S^q$), whose dynamics are governed by the following equations:

$$\frac{dS^n}{dt} = -\lambda^b S^n\left(1+(\eta-1)\frac{S^n}{N^n}\right)+\frac{S^I}{d^{hh}}+\frac{S^q}{d^q}$$

$$\frac{dS^E}{dt} = \lambda^b S^n(\eta-1)\frac{S^n}{N^n}-\lambda^b S^E-\frac{S^E}{d^E}$$

$$\frac{dS^I}{dt} = \frac{S^E}{d^E}-\left(\lambda^{sa}+\lambda^{ss}+\lambda^t+\lambda^b\right)S^I-(\eta-1)\chi^q\left(\frac{I_p^f}{d^p}\frac{S^I}{N^{fq}}+\frac{I_p^{sa}}{d^p}\frac{S^I}{N^{sq}}\right)-\frac{S^I}{d^{hh}} \tag{S.1}$$

$$\frac{dS^q}{dt} = -\lambda^q S^q+(\eta-1)\chi^q\left(\frac{I_p^f}{d^p}\frac{S^I}{N^{fq}}+\frac{I_p^{sa}}{d^p}\frac{S^I}{N^{sq}}\right)-\frac{S^q}{d^q}$$

Here $S^n$ includes all those susceptible individuals that are exposed to risk of transmission from individuals outside of their household (with force of infection $\lambda^b$ (equation (S.3))), but who have no infected (latent or otherwise) individuals within their households. Each time an individual from $S^n$ becomes latently infected, $\eta-1$ additional individuals from the sub-population $N^n$, which includes all those individuals in households with no infection (equation (S.9)), are identified as being from the same household as the new latent individual by moving them to a new category; in the case of household members from $S^n$, this new category is $S^E$. Movement from $S^E$ to $S^I$ happens at the rate $1/d^E$ where $d^E$ is the average duration of an individual in the latent state. $S^E$ and $S^I$ are exposed to between-household transmission with force of infection $\lambda^b$, and individuals in $S^I$ are additionally exposed to within-household transmission with force of infection $\left(\lambda^{sa}+\lambda^{ss}+\lambda^t\right)$ (equation (S.3)). Individuals in $S^I$ either progress to the latent infection state through within-household transmission or return to $S^n$ at rate $1/d^{hh}$, where $d^{hh}$ is the mean duration that a household is expected to remain infectious. We estimated $d^{hh}$ by running a stochastic version of the SEIR dynamics 50,000 times within a household of size $\eta$ with the within-household transmission rates (equation (S.4)) using the ssar package in R [1]. By calculating the mean time at which no infectious individuals remained in the household over the simulations, we estimate $d^{hh} = 10.56$. Using the appropriate subsets of the simulations we also estimated the mean time a household remains infectious if the first infectious individual is asymptomatic $d^{hha} = 9.87$ and if the first infectious individual is symptomatic $d^{hhs} = 12.16$, which are used in equation (S.9). A household where a pre-symptomatic person who was the first infection in their household becomes symptomatic has a probability $\chi^q$ of going into quarantine, calculated as:

$$\chi^q = \begin{cases} 0 & \text{if } t < t^{q1} \mid t \geq t^{q2} \\ \min\left(\hat{\chi}^q, \hat{\chi}^q\left(\frac{t - t^{q1}}{\tau^q}\right)\right) & \text{if } t^{q1} \leq \mathrm{t} < t^{q2} \end{cases} \tag{S.2}$$

where $t^{q1}$ and $t^{q2}$ are the start and end times of the implementation period, and $\tau^q$ is the length of the scale-up period, during which the proportion of households that comply with quarantine increases linearly, and after which compliance remains constant at $\hat{\chi}^q$ until $t^{q2}$. Households where a pre-symptomatic person becomes symptomatic after being infected by an asymptomatic first infection in the same household also enter quarantine, again with probability $\chi^q$. These two types of quarantining event occur according to $\chi^q I_p^f / d^p$ and $\chi^q I_p^{sa} / d^p$ respectively, where $d^p$ is the mean duration in the pre-symptomatic disease state, $I_p^f$ are pre-symptomatic first infections in their household, and $I_p^{sa}$ are pre-symptomatic infections that are secondary to an asymptomatic first infection in their household. When the first type of quarantine event occurs $\eta - 1$ other individuals, representing the household of the newly symptomatic individual, move into quarantine with them from the sub-population $N^{fq}$ (equation (S.9)), while in the second type of quarantine event the $\eta - 1$ household members come from sub-population $N^{sq}$ (equation (S.9)). Individuals from $S^I$ that form part of newly quarantining households move to category $S^q$, where they remain until they are infected by within-household transmission (with force of infection $\lambda^q$; equation (S.3)) or until the quarantine period of duration $d^q$ ends and they return to $S^n$.

The forces of infection in equation (S.1) are: 1) $\lambda^b$, which represents between-household transmission (equations (S.4, S.5, S.7)) and targets those in non-quarantined households; 2) $\lambda^t$, which creates tertiary, quaternary, etc. cases within households through transmission from cases in the second generation onwards; 3) $\lambda^{sa}$, which creates second generation cases in households where the first infection was asymptomatic; 4) $\lambda^{ss}$, which creates second generation cases in households where the first infection was symptomatic; and 5) $\lambda^q$, which targets quarantined individuals with within-household transmission. These forces of infection are defined:

$$\lambda^b = \frac{\beta_a^{bhh}\left(I_a^f + I_a^s + I_a^{sa} + I_a^b\right) + \beta_p^{bhh}\left(I_p^f + I_p^s + I_p^{sa} + I_p^b\right) + \beta_s^{bhh}\left(I_s^f + I_s^s + I_s^b\right)}{N}$$

$$\lambda^t = \frac{\beta_a^{hh}\left(I_a^s + I_a^{sa} + I_a^b\right) + \beta_p^{hh}\left(I_p^s + I_p^{sa} + I_p^b\right) + \beta_s^{hh}\left(I_s^s + I_s^b\right)}{N^I}$$

$$\lambda^{sa} = \frac{\beta_a^{hh} I_a^f}{N^I} \tag{S.3}$$

$$\lambda^{ss} = \frac{\beta_p^{hh} I_p^f + \beta_s^{hh} I_s^f}{N^I}$$

$$\lambda^q = \frac{\beta_a^{hh}\left(I_a^q + I_a^{qa}\right) + \beta_p^{hh}\left(I_p^q + I_p^{qp}\right) + \beta_s^{hh}\left(I_s^q + I_s^{qs}\right)}{N^q}$$

where $\beta_a^{bhh}$, $\beta_p^{bhh}$ and $\beta_s^{bhh}$ are the between-household transmission rates, and $\beta_a^{hh}$, $\beta_p^{hh}$ and $\beta_s^{hh}$ the within-household transmission rates, for each of the three infectious states. These between- and within- household rates were estimated by breaking down the overall transmission rates using the household secondary attack rate, $\sigma = 0.166$ [2], and the mean household size in Bangladesh, $\eta = 4$ [3], as follows (taking the transmission rate of asymptomatic cases $\beta_a$ as an example):

$$\beta_a = \beta_a^{hh} + \beta_a^{bhh}$$

$$\beta_a^{hh} = \frac{\sigma(\eta - 1)}{R_0}\beta_a \tag{S.4}$$

$$\hat{\beta}_a^{bhh} = \left(1 - \frac{\sigma(\eta - 1)}{R_0}\right)\beta_a$$

Here, $\beta_a^{hh}$ is the within-household transmission rate for asymptomatic individuals, while $\hat{\beta}_a^{bhh}$ is the between-household transmission rate prior to accounting for the effects of lockdown and masks.

During lockdown, we assume that the between-household transmission rates are reduced by the proportion $\varepsilon^{ld}$ for those that are compliant (a proportion $\chi^{ld}$ of the population), while transmission rates of the non-compliant and of non-symptomatic essential workers are unaffected. Symptomatic essential workers, who are assumed to be unable to work due to illness, are treated in the same way as the rest of the population, with only a proportion $\chi^{ld}$ complying with lockdown and having reduced transmission. The following between-household transmission rates can then be defined:

$$\beta_a^{bhh} = \hat{\beta}_a^{bhh}\left(f^{ldw} + \left(1 - f^{ldw}\right)\left(\left(1 - \chi^{ld}\right) + \left(1 - \varepsilon^{ld}\right)\chi^{ld}\right)\right)$$

$$\beta_p^{bhh} = \hat{\beta}_p^{bhh}\left(f^{ldw} + \left(1 - f^{ldw}\right)\left(\left(1 - \chi^{ld}\right) + \left(1 - \varepsilon^{ld}\right)\chi^{ld}\right)\right) \tag{S.5}$$

$$\beta_s^{bhh} = \hat{\beta}_s^{bhh}\left(\left(1 - \chi^{ld}\right) + \left(1 - \varepsilon^{ld}\right)\chi^{ld}\right)$$

The lockdown period starts at time $t^{ld1}$ and ends at time $t^{ld2}$, with compliance being zero outside of this period. A scale-up period of length $\tau^{ld}$, starting at $t^{ld1}$, was implemented, during which compliance increased linearly from zero to $\hat{\chi}^{ld}$. Following the scale-up period, compliance declines exponentially at rate $r^{ld}$ towards a minimum of $\hat{\chi}_{\min}^{ld}$. $\chi^{ld}$ is, therefore, defined by:

$$\chi^{ld} = \begin{cases} 0 & \text{if } t < t^{ld1} \mid t \geq t^{ld2} \\ \hat{\chi}^{ld}\left(\dfrac{t - t^{ld1}}{\tau^{ld}}\right) & \text{if } t^{ld1} \leq \mathrm{t} < \left(t^{ld1} + \tau^{ld}\right) \\ e^{-r^{ld}\left(t - t^{ld1} - \tau^{ld}\right)\left(\hat{\chi}^{ld} - \hat{\chi}^{ld}_{\min}\right)} + \hat{\chi}^{ld}_{\min} & \text{if } \left(t^{ld1} + \tau^{ld}\right) \leq \mathrm{t} < t^{ld2} \end{cases} \tag{S.6}$$

Masks are assumed to block a proportion, $\varepsilon^m$, of between-household transmission from compliant individuals, while also blocking a proportion, $\varepsilon^m \rho^m$, of transmission to compliant individuals, where $0 \leq \rho^m \leq 1$. When incorporating the effect of masks in addition to the effect of lockdown, the overall between-household transmission rates become:

$$\begin{aligned} \beta_a^{bhh} &= \hat{\beta}_a^{bhh}\left(f^{ldw} + \left(1 - f^{ldw}\right)\left(\left(1 - \chi^{ld}\right) + \left(1 - \varepsilon^{ld}\right)\chi^{ld}\right)\right)\left(\left(1 - \varepsilon^m \chi^m\right)\left(1 - \varepsilon^m \rho^m \chi^m\right)\right) \\ \beta_p^{bhh} &= \hat{\beta}_p^{bhh}\left(f^{ldw} + \left(1 - f^{ldw}\right)\left(\left(1 - \chi^{ld}\right) + \left(1 - \varepsilon^{ld}\right)\chi^{ld}\right)\right)\left(\left(1 - \varepsilon^m \chi^m\right)\left(1 - \varepsilon^m \rho^m \chi^m\right)\right) \\ \beta_s^{bhh} &= \hat{\beta}_s^{bhh}\left(\left(1 - \chi^{ld}\right) + \left(1 - \varepsilon^{ld}\right)\chi^{ld}\right)\left(\left(1 - \varepsilon^m \chi^m\right)\left(1 - \varepsilon^m \rho^m \chi^m\right)\right) \end{aligned} \tag{S.7}$$

Here, $\chi^m$ is the proportion of people that are compliant with mask wearing, which is calculated through time as:

$$\chi^m = \begin{cases} 0 & \text{if } t < t^{m1} \mid t \geq t^{m2} \\ \min\left(\hat{\chi}^m, \hat{\chi}^m\left(\dfrac{t - t^{m1}}{\tau^m}\right)\right) & \text{if } t^{m1} \leq \mathrm{t} < t^{m2} \end{cases} \tag{S.8}$$

where $t^{m1}$ and $t^{m2}$ are the start and end times of the compulsory mask wearing period, and $\tau^m$ is the length of the scale-up period, during which compliance increases linearly to $\hat{\chi}^m$.

In equations (S.1, S.3), and in the following equations, a number of sub-populations of the total population $N$ that are composed of individuals in multiple model compartments are referred to. These sub-populations are defined as follows:

$$\begin{aligned} N^n &= S^n + R^n \\ N^I &= S^I + E^{ss} + E^{sa} + E^t + E^b + I_a^f + I_a^s + I_a^{sa} + I_a^b + I_p^f + I_p^s + I_p^{sa} + I_p^b + I_s^f + I_s^s + I_s^b + R^I + R^{I_a^f} + R^{I_s^f} + R^{I^b} \\ N^q &= S^q + E^q + E^{qE} + I_a^q + I_a^{qa} + I_p^q + I_p^{qp} + I_s^q + I_s^{qs} + R^{qR} + R^{I_a^{qa}} + R^{I_s^{qs}} + R^{I_a^q} + R^{I_s^q} \\ N^{fq} &= S^I + E^{ss} + E^b + I_p^b + I_a^b + R^I \\ N^{sq} &= S^I + E^{sa} + E^b + I_p^{sa} + I_p^b + I_a^f + I_a^{sa} + I_a^b + R^I + R^{I_a^f} \end{aligned} \tag{S.9}$$

We divide the population that is latently infected with SARS-CoV-2 infection (denoted $E$ ) into seven sub-categories ( $E = E^f + E^b + E^{ss} + E^{sa} + E^t + E^q + E^{qE}$ ), with dynamics as follows:

$$\frac{dE^f}{dt} = \lambda^b S^n - \frac{E^f}{d^E}$$

$$\frac{dE^b}{dt} = \lambda^b \left(S^E + S^I\right) - \frac{E^b}{d^E} - (\eta - 1)\chi^q \left( \frac{I_p^f}{d^p} \frac{E^b}{N^{fq}} + \frac{I_p^{sa}}{d^p} \frac{E^b}{N^{sq}} \right)$$

$$\frac{dE^{ss}}{dt} = \lambda^{ss} S^I - \frac{E^{ss}}{d^E} - (\eta - 1)\chi^q \frac{I_p^f}{d^p} \frac{E^{ss}}{N^{fq}}$$

$$\frac{dE^{sa}}{dt} = \lambda^{sa} S^I - \frac{E^{sa}}{d^E} - (\eta - 1)\chi^q \frac{I_p^{sa}}{d^p} \frac{E^{sa}}{N^{sq}}$$

$$\frac{dE^t}{dt} = \lambda^t S^I - \frac{E^t}{d^E}$$

$$\frac{dE^q}{dt} = \lambda^q S^q - \frac{E^q}{d^E}$$

$$\frac{dE^{qE}}{dt} = (\eta - 1)\chi^q \left( \frac{I_p^f}{d^p} \frac{\left(E^{ss} + E^b\right)}{N^{fq}} + \frac{I_p^{sa}}{d^p} \frac{\left(E^{sa} + E^b\right)}{N^{sq}} \right) - \frac{2E^{qE}}{d^E} \quad \text{(S.10)}$$

Here $1/d^E$ is the rate at which latently infected individuals move on to the infectious disease states. Individuals that are quarantined while exposed, $E^{qE}$, could have been at any stage in their latent period at the time of quarantine, so we assume that they progress at twice the rate.

The population that is asymptomatically infectious ( $I_a$ ) is divided into six sub-categories $\left(I_a = I_a^f + I_a^b + I_a^s + I_a^{sa} + I_a^q + I_a^{qa}\right)$:

$$\frac{dI_a^f}{dt} = \frac{f^a E^f}{d^E} - \frac{I_a^f}{d^a} - (\eta - 1)\chi^q \frac{I_p^{sa}}{d^p}\frac{I_a^{sa}}{N^{sq}}$$

$$\frac{dI_a^b}{dt} = \frac{f^a E^b}{d^E} - \frac{I_a^b}{d^a} - (\eta - 1)\chi^q \left(\frac{I_p^f}{d^p}\frac{I_a^b}{N^{fq}} + \frac{I_p^{sa}}{d^p}\frac{I_a^b}{N^{sq}}\right)$$

$$\frac{dI_a^s}{dt} = \frac{f^a\left(E^{ss} + E^t\right)}{d^E} - \frac{I_a^s}{d^a}$$

$$\frac{dI_a^{sa}}{dt} = \frac{f^a E^{sa}}{d^E} - \frac{I_a^{sa}}{d^a} - (\eta - 1)\chi^q \frac{I_p^{sa}}{d^p}\frac{I_a^{sa}}{N^{sq}}$$

$$\frac{dI_a^q}{dt} = \frac{f^a\left(E^q + 2E^{qE}\right)}{d^E} - \frac{I_a^q}{d^a}$$

$$\frac{dI_a^{qa}}{dt} = (\eta - 1)\chi^q \left(\frac{I_p^f}{d^p}\frac{I_a^b}{N^{fq}} + \frac{I_p^{sa}}{d^p}\frac{I_a^f + I_a^b + I_a^{sa}}{N^{sq}}\right) - \frac{2I_a^{qa}}{d^a} \quad \text{(S.11)}$$

The number of people entering the first four asymptomatic sub-categories is a fraction $f^a$ of those leaving the latently infected categories. Asymptomatic people recover at the rate $1/d^a$ . Those that are quarantined from the $I_a^b$ and $I_a^{sa}$ categories may have been quarantined in any stage of their infectious period, and those in $I_a^{qa}$ , therefore, are assumed to remain there on average for half the mean asymptomatic duration .

Pre-symptomatic individuals are also divided into six sub-categories $\left(I_p = I_p^f + I_p^b + I_p^s + I_p^{sa} + I_p^q + I_p^{qp}\right)$:

$$\frac{dI_p^f}{dt} = \frac{\left(1-f^a\right)E^f}{d^E} - \frac{I_p^f}{d^p}$$

$$\frac{dI_p^b}{dt} = \frac{\left(1-f^a\right)E^b}{d^E} - \frac{I_p^b}{d^p} - (\eta-1)\chi^q\left(\frac{I_p^f}{d^p}\frac{I_p^b}{N^{fq}} + \frac{I_p^{sa}}{d^p}\frac{I_p^b}{N^{sq}}\right)$$

$$\frac{dI_p^s}{dt} = \frac{\left(1-f^a\right)\left(E^{ss}+E^t\right)}{d^E} - \frac{I_p^s}{d^p}$$

$$\frac{dI_p^{sa}}{dt} = \frac{\left(1-f^a\right)E^{sa}}{d^E} - \frac{I_p^{sa}}{d^p} - (\eta-1)\chi^q\frac{I_p^{sa}}{d^p}\frac{I_p^{sa}}{N^{sq}}$$

$$\frac{dI_p^q}{dt} = \frac{\left(1-f^a\right)\left(E^q+2E^{qE}\right)}{d^E} - \frac{I_p^q}{d^p}$$

$$\frac{dI_p^{qp}}{dt} = (\eta-1)\chi^q\left(\frac{I_p^f}{d^p}\frac{I_p^b}{N^{fq}} + \frac{I_p^{sa}}{d^p}\frac{I_p^b+I_p^{sa}}{N^{sq}}\right) - \frac{2I_p^{qp}}{d^p} \qquad \text{(S.12)}$$

The proportion of latently infected individuals that do not become asymptomatic, $1-f^a$, enter the first five of these pre-symptomatic categories. They then move on to the symptomatic categories at the rate $1/d^p$. Those that are quarantined from the $I_p^b$ and $I_p^{sa}$ categories could be at any point in their pre-symptomatic period, and are therefore assumed to remain in $I_p^{qp}$ for half the usual period.

The model has five symptomatic sub-categories $\left(I_s = I_s^f + I_s^b + I_s^s + I_s^q + I_s^{qs}\right)$ with dynamics:

$$\frac{dI_s^f}{dt} = \frac{\left(1-\chi^q\right)I_p^f}{d^p} - \frac{I_s^f}{d^s}$$

$$\frac{dI_s^b}{dt} = \frac{I_p^b}{d^p} - \frac{I_s^b}{d^s}$$

$$\frac{dI_s^s}{dt} = \frac{\left(1-\chi^q\right)I_p^{sa} + I_p^s}{d^p} - \frac{I_s^s}{d^s} \tag{S.13}$$

$$\frac{dI_s^q}{dt} = \frac{I_p^q + 2I_p^{qp}}{d^p} - \frac{I_s^q}{d^s}$$

$$\frac{dI_s^{qs}}{dt} = \frac{\chi^q\left(I_p^f + I_p^{sa}\right)}{d^p} - \frac{I_s^{qs}}{d^s}$$

Pre-symptomatic individuals that were the first infection in their household either enter $I_s^f$ if not complying with a quarantine (with probability $\left(1-\chi^q\right)$) or enter the quarantined category $I_s^{qs}$. Those that were secondary to an asymptomatic first household infection either enter $I_s^s$ if not quarantining or $I_s^{qs}$ if quarantining. Symptomatic individuals are removed at rate $1/d^s$.

There are eleven sub-categories for removed individuals within the model $\left(R = R^n + R^E + R^I + R^{I_a^f} + R^{I_s^f} + R^{I^b} + R^{qR} + R^{I_a^{qa}} + R^{I_s^{qs}} + R^{I_a^q} + R^{I_s^q}\right)$:

$$
\begin{aligned}
\frac{dR^n}{dt} &= \frac{I_a^s + I_a^{sa}}{d^a} + \frac{I_s^s}{d^s} + \frac{R^I}{d^{hh}} + \frac{R^{qR}}{d^q} + \frac{R^{I_a^f}}{d^{hha} - d^a} + \frac{R^{I_s^f}}{d^{hhs} - d^p - d^s} + \frac{2R^{I^b}}{d^{hh} - \left(\left(1 - f^a\right)\left(d^p + d^s\right) + f^a d^a\right)} \\
&\quad + \frac{R^{I_a^{qa}}}{d^q - d^a/2} + \frac{2R^{I_a^q}}{d^q - d^a} + \frac{R^{I_s^{qs}} + 2R^{I_s^q}}{d^q - d^s} - (\eta - 1)\lambda^b S^n \frac{R^n}{N^n}
\end{aligned}
$$

$$\frac{dR^E}{dt} = (\eta - 1)\lambda^b S^n \frac{R^n}{N^n} - \frac{R^E}{d^E}$$

$$\frac{dR^I}{dt} = \frac{R^E}{d^E} - \frac{R^I}{d^{hh}} - (\eta - 1)\chi^q \left(\frac{I_p^f}{d^p}\frac{R^I}{N^{fq}} + \frac{I_p^{sa}}{d^p}\frac{R^I}{N^{sq}}\right)$$

$$\frac{dR^{I_a^f}}{dt} = \frac{I_a^f}{d^a} - \frac{R^{I_a^f}}{d^{hha} - d^a} - (\eta - 1)\chi^q \frac{I_p^{sa}}{d^p}\frac{R^{I_a^f}}{N^{sq}}$$

$$\frac{dR^{I_s^f}}{dt} = \frac{I_s^f}{d^s} - \frac{R^{I_s^f}}{d^{hha} - d^p - d^s}$$

$$\frac{dR^{I^b}}{dt} = \frac{I_a^b}{d^a} + \frac{I_s^b}{d^s} - \frac{2R^{I^b}}{d^{hh} - \left(\left(1 - f^a\right)\left(d^p + d^s\right) + f^a d^a\right)}$$

$$\frac{dR^{qR}}{dt} = (\eta - 1)\chi^q \left(\frac{I_p^f}{d^p}\frac{R^I}{N^{fq}} + \frac{I_p^{sa}}{d^p}\frac{\left(R^I + R^{I_a^f}\right)}{N^{sq}}\right) - \frac{R^{qR}}{d^q}$$

$$\frac{dR^{I_a^{qa}}}{dt} = \frac{2I_a^{qa}}{d^a} - \frac{R^{I_a^{qa}}}{d^q - d^a/2}$$

$$\frac{dR^{I_s^{qs}}}{dt} = \frac{I_s^{qs}}{d^s} - \frac{R^{I_s^{qs}}}{d^q - d^s}$$

$$\frac{dR^{I_a^q}}{dt} = \frac{I_a^q}{d^a} - \frac{2R^{I_a^q}}{d^q - d^a} \qquad \text{(S.14)}$$

$$\frac{dR^{I_s^q}}{dt} = \frac{I_s^q}{d^s} - \frac{2R^{I_s^q}}{d^q - d^s}$$

$R^n$ includes all removed individuals that are not exposed to within-household transmission. When a new individual enters $E^f$, household members of that individual that are drawn from $R^n$ move to $R^E$, where they remain until they join $R^I$ after the latently infected household member becomes infectious. Those in $R^I$ remain there until the household is cleared of infection (at rate $1/d^{hh}$) or until they are quarantined and enter $R^{qR}$. Asymptomatic and symptomatic individuals who were the first infection in their household enter categories $R^{I_a^f}$ and $R^{I_s^f}$ respectively for the remainder of their household's infectious period, after which they join $R^n$. Infectious individuals that were infected by between-household transmission after the first infection in their household enter $R^{I^b}$ and remain there for $d^{hh}$ minus the average time an individual is infectious. Infectious non-quarantined individuals that were infected by within-household transmission move directly to $R^n$ on removal. Those that were quarantined when they became symptomatic or as asymptomatic individuals join $R^{I_a^{qa}}$ and $R^{I_s^{qs}}$ respectively for the remainder of the quarantine period, while those that were quarantined in earlier disease states join $R^{I_a^q}$ and $R^{I_s^q}$ until quarantine ends.

The equations governing the health outcome categories (Fig. 1) are defined:

$$\frac{dD_{wait}}{dt} = f^D\left(\frac{I_p^f + I_p^b + I_p^s + I_p^{sa} + I_p^q + 2I_p^{qp}}{d^p}\right) - \frac{D_{wait}}{d^{D_{wait}}}$$

$$\frac{dD}{dt} = \frac{D_{wait}}{d^{D_{wait}}}$$

$$\frac{dH_{wait}}{dt} = f^H\left(1 - f^{ICU}\right)\left(\frac{I_p^f + I_p^b + I_p^s + I_p^{sa} + I_p^q + 2I_p^{qp}}{d^p}\right) - \frac{H_{wait}}{d^{H_{wait}}}$$

$$\frac{dH}{dt} = \frac{H_{wait}}{d^{H_{wait}}} - \frac{H}{d^H}$$

$$\frac{dICU_{wait}}{dt} = f^H f^{ICU}\left(\frac{I_p^f + I_p^b + I_p^s + I_p^{sa} + I_p^q + 2I_p^{qp}}{d^p}\right) - \frac{ICU_{wait}}{d^{H_{wait}}}$$

$$\frac{dICU}{dt} = \frac{ICU_{wait}}{d^{H_{wait}}} - \frac{ICU}{d^{ICU}}$$

$$\frac{dRecup}{dt} = \left(1 - \frac{f^D}{f^H}\right)\left(\frac{H}{d^H} + \frac{ICU}{d^{ICU}}\right) - \frac{Recup}{d^{Recup}} \qquad \text{(S.15)}$$

Working days lost on each day of the model simulations are estimated as follows:

$$
\begin{aligned}
W = & \frac{5}{7}\left( \left(D+Recup\right)\frac{N-N^q}{N} + I_s + \left(N^q - I_s^q - I_s^{qs}\right) \right) f^e \\
& + \frac{5}{7}\chi^{ld} \left( \begin{array}{l} \left(S-S^q\right) + \left(E-E^q-E^{qE}\right) + \left(I_p - I_p^q - I_p^{qp}\right) + \left(I_a - I_a^q - I_a^{qa}\right) \\ + \left( \left(R - R^{qR} - R^{I_a^{qa}} - R^{I_s^{qs}} - R^{I_a^q} - R^{I_s^q}\right)\left(1 - \frac{D+Recup}{R}\right) \right) \end{array} \right) \left(f^e - f^{ldw}\right) \\
& + \frac{5}{7}\left( \left(D+Recup\right)\frac{N-N^q}{N} + \frac{I_s N^I}{N^I + N^q} \right)\left( \left(f^e - f^{ldw}\right)\left(1-\chi^{ld}\right) + f^{ldw} \right) f^c \\
& + \frac{5}{7} G\left(\eta - 1 - f^c\right)\left( \left(f^e - f^{ldw}\right)\left(1-\chi^{ld}\right) + f^{ldw} \right)\left(1 - \frac{Recup+D}{N}\right) \\
& + \frac{5}{7}\frac{v\chi^q\left(N-N^q\right)}{365}\left(1 - \frac{I_s^f + I_s^b + I_s^s}{N-N^q} - \frac{Recup+D}{N}\right)\left( \left(f^e - f^{ldw}\right)\left(1-\chi^{ld}\right) + f^{ldw} \right)
\end{aligned}
\qquad \text{(S.16)}
$$

The first line in this equation describes working days lost by employed individuals dying, being sick or being under quarantine. Additional days lost by (non-symptomatic) employed people who are not essential workers during lockdown are accounted for in the second line. The third line deals with days lost by employed people who stay off work to take over caring responsibilities from those working within the household. These additional working days lost to replace household workers only occur if the household is not already quarantining, or if the employed person replacing the household member is not already off work due to lockdown, as indicated by multiplication by $\left(f^e - f^{ldw}\right)\left(1-\chi^{ld}\right) + f^{ldw}$ rather than simply $f^e$. The fourth line of equation (S.16) describes working days lost by those grieving people who have died due to COVID-19. Here $G$ is the number of people who have died recently being grieved. The number of deaths being grieved on day $d$ is estimated as:

$$G(d) = D(d) - D(d-g) \qquad \text{(S.17)}$$

where $D(d)$ is the number of deaths on day $d$ and $g$ is the mean number of days for which the death is grieved. The number of grievers for each death is assumed to equal the household size minus one. The grievers are assumed not to lose working days if they are already off work due to lockdown, as described by multiplication by $\left(f^e - f^{ldw}\right)\left(1-\chi^{ld}\right) + f^{ldw}$ or if they are already filling in for the caring responsibilities of the person being grieved (by subtracting $f^c$ from the number of grievers). The final line of equation (S.16) refers to working days lost due to quarantining of households due to influenza-like illnesses (ILIs) that are not COVID-19. These households come from the population that is not already quarantining, $N^n + N^I$. The proportion of this population that is dead, recuperating or symptomatic with COVID-19 is assumed not to lose any further working days, as the proportion that already under lockdown. The compliance of households with other influenza-like illnesses with quarantine is assumed to be equal to that for those with COVID-19. Note that there may be some overestimation of working days lost in household quarantine scenarios since we don’t account for working days already lost by workers taking over duties from ill or dead home workers when estimating days lost due to quarantine associated with ILIs.

### Supplement B: Supplementary Tables

**Table S1: Age-distribution of Dhaka District.** Counts of the district population in each age category are taken from the 2011 census of Bangladesh[4]. The proportion of the population in each category is used to inform the proportions of: 1) infections that are asymptomatic $f^a$, 2) symptomatic infections that lead to death $f^D$, and 3) symptomatic infections that are hospitalised $f^H$.

| Age group | Count | Proportion of population |
|---|---|---|
| 0-9 | 2106777 | 0.175 |
| 10-14 | 2387793 | 0.198 |
| 20-29 | 3095980 | 0.257 |
| 30-39 | 2004910 | 0.166 |
| 40-49 | 1230235 | 0.102 |
| 50-59 | 656006 | 0.054 |
| 60-69 | 351786 | 0.029 |
| 70-79 | 145844 | 0.012 |

**Table S2: Age-dependent severity of infection.** Estimates from literature of the percentage of SARS-CoV-2 infections that are fatal [5], the percentage of symptomatic cases that are hospitalised [5], and the percentage of infections that are symptomatic [6] for each of nine ten-year age bands.

| Age group | % cases that are fatal | % symptomatic cases that are hospitalised | % cases that are symptomatic |
|---|---|---|---|
| 0-9 | 0 | 0 | 0.147372 |
| 10-19 | 0.09 | 0.8 | 0.073029 |
| 20-29 | 0.1 | 0.8 | 0.296123 |
| 30-39 | 0.12 | 1 | 0.419104 |
| 40-49 | 0.23 | 1.9 | 0.444587 |
| 50-59 | 0.68 | 5.4 | 0.563572 |
| 60-69 | 1.87 | 15.1 | 0.816944 |
| 70-79 | 4.14 | 33.3 | 0.75056 |
| 80-89 | 7.68 | 61.8 | 0.75056 |

**Table S3: Descriptions of all model state variables**

| State variable | Description |
|---|---|
| $S^n$ | Susceptible individuals that are not in an infected or quarantined household |
| $S^E$ | Susceptible individuals who are in a household that does not yet have any infectious individuals, but that has a latently infected individual |
| $S^I$ | Susceptible individuals that are in a household that includes one or more infectious individuals, but is not under quarantine |
| $S^q$ | Susceptible individuals that are in a household that has been quarantined |
| $E^f$ | Latently infected individuals that are the first infected in their household |
| $E^{ss}$ | Latent individuals that are in the second generation of cases in a non-quarantined household where the first infection followed the pre-symptomatic->symptomatic progression |
| $E^b$ | Latent individuals that were infected by between-household transmission when there was already an infected individual in their household |
| $E^{sa}$ | Latent individuals that are in the second generation of cases in a non-quarantined household where the first infection was asymptomatic |
| $E^t$ | Latent individuals that are in the third (or later) generation of cases in a non-quarantined household |
| $E^q$ | Quarantined latent individuals that were first quarantined while they were still susceptible |
| $E^{qE}$ | Quarantined latent individuals that were first quarantined when they were already in the latent state |
| $I_a^f$ | Asymptomatic individuals that are not in a quarantined household and are the first infection in their household |
| $I_a^b$ | Asymptomatic individuals that were infected by between-household transmission when there was already an infected individual in their household |
| $I_a^{sa}$ | Asymptomatic individuals that are not in a quarantined household and are secondary to a case in $I_a^f$ |
| $I_a^s$ | Asymptomatic individuals that are in a non-quarantined household and are secondary to the first infection in the household if that infection was symptomatic ( $I_p^f$ or $I_s^f$ ) or in the third (or later) generation of cases in the household |
| $I_a^q$ | Quarantined asymptomatic individuals that were first quarantined while they were in an earlier disease state |
| $I_a^{qa}$ | Quarantined asymptomatic individuals that were first quarantined while already asymptomatic |
| $I_p^f$ | Pre-symptomatic individuals that are not in a quarantined household and are the first in their household to become infectious |
| $I_p^b$ | Pre-symptomatic individuals that were infected by between-household transmission when there was already an infected individual in their household |
| $I_p^{sa}$ | Pre-symptomatic individuals that are not in a quarantined household and are secondary to a case in $I_a^f$ |
| $I_p^s$ | Pre-symptomatic individuals that are in a non-quarantined household and are secondary to the first infection in the household if that infection was symptomatic ( $I_p^f$ or $I_s^f$ ) or in the third (or later) generation of cases in the household |
| $I_p^q$ | Quarantined pre-symptomatic individuals that were first quarantined while they were in an earlier disease state |
| $I_p^{qp}$ | Quarantined pre-symptomatic individuals that were first quarantined while |

| | |
|---|---|
| | already asymptomatic |
| $I_s^f$ | Symptomatic individuals that are not in a quarantined household and are the first in their household to become infectious. |
| $I_s^b$ | Symptomatic individuals that were infected by between-household transmission when there was already an infected individual in their household |
| $I_s^s$ | Symptomatic individuals that are not in a quarantined household and are not the first in their household to become infectious. |
| $I_s^q$ | Quarantined symptomatic individuals that were first quarantined while they were in an earlier disease state |
| $I_s^{qs}$ | Quarantined symptomatic individuals that were first quarantined while already asymptomatic |
| $R^n$ | Removed individuals whose household is not currently infected, but was at an earlier point in time |
| $R^E$ | Removed individuals who are in a household that does not yet have any infectious individuals, but that has a latently infected individual |
| $R^I$ | Removed individuals who are in an infected non-quarantined household and were already removed at the beginning of the household's current outbreak |
| $R^{I_a^f}$ | Individuals that were removed from $I_a^f$ in an ongoing household outbreak |
| $R^{I_s^f}$ | Individuals that were removed from $I_s^f$ in an ongoing household outbreak |
| $R^{I^b}$ | Individuals that were removed from $I_a^b$ or $I_s^b$ in an ongoing household outbreak |
| $R^{qR}$ | Removed individuals who are in a quarantined household and were already removed at the beginning of the household's current outbreak |
| $R^{I_a^{qa}}$ | Individuals that were removed from $I_a^{qa}$ in an ongoing household outbreak and that remain under quarantine |
| $R^{I_s^{qs}}$ | Individuals that were removed from $I_s^{qs}$ in an ongoing household outbreak and that remain under quarantine |
| $R^{I_a^q}$ | Individuals that were removed from $I_a^q$ in an ongoing household outbreak and that remain under quarantine |
| $R^{I_s^q}$ | Individuals that were removed from $I_s^q$ in an ongoing household outbreak and that remain under quarantine |
| $D_{wait}$ | Holding category for individuals who will die of COVID-19 |
| $D$ | Total number of deaths due to COVID-19 |
| $H_{wait}$ | Holding category for individuals who will require a non-ICU hospital bed |
| $H$ | Total number of individuals who are in a non-ICU hospital bed |
| $ICU_{wait}$ | Holding category for individuals who will require an ICU hospital bed |
| $ICU$ | Total number of individuals who are in an ICU hospital bed |
| $Recup$ | Total number of individuals who are recuperating following hospital discharge |
| $N$ | The sum of all individuals in the categories described in equations (S.1,10-14). Equal to the population size – see Table S4 for the assumed value |
| $N^n$ | The total number of individuals that are not in infected household and are exposed to between-household transmission |
| $N^I$ | The total number of individuals in non-quarantined infected households, who are exposed to within-household transmission |
| $N^q$ | The total number of individuals in quarantined infected households, who are exposed to within-household transmission |
| $N^{fq}$ | The sub-population from which quarantining household members of newly |

| | |
|---|---|
| | symptomatic $I_p^f \to I_s^{qs}$ are drawn |
| $N^{sq}$ | The sub-population from which quarantining household members of newly symptomatic $I_p^{sa} \to I_s^{qs}$ are drawn |

**Table S4: Values, descriptions and sources of all model parameters.** For those parameters for which a bracketed range is provided, this is the range of possible values of those parameters considered during sensitivity analyses.

| Parameter | Value | Description | Source |
|---|---|---|---|
| $t^{intro}$ | 15/02/2020 (13/01/2020-08/03/2020) | The date on which the first infectious cases are introduced to the modelled population. | The first three cases of COVID-19 in Bangladesh were confirmed on 08/03/2020, but it is likely the disease had started circulating undetected prior to this, with estimates from genomic data indicating an introduction in mid-February [7]. At least 8 introductions from three different lineages are estimated to have occurred prior to the ban on international travel (March 21st) [7]; we therefore initialise the model with 8 cases at $t^{intro}$. 13/01/2020 was the date of the first detection of COVID-19 outside China and is assumed to be the earliest possible introduction date. |
| $R_0$ | 3.57 (1.56-5.7) | The basic reproduction number: the expected number of secondary cases generated by a single infectious individual in a fully susceptible population. $R_0 = f^a \beta^a d^a + \left(1 - f^a\right)\left(\beta^p d^p + \beta^s d^s\right)$. | Tuned to match ECDC (European Centre for Disease Prevention and Control) data for Bangladesh [8]. Minimum potential value is taken from [9] and maximum is the median estimate from [10] |
| $\beta_a$ | 0.40 | Daily transmission rate of asymptomatic cases. This rate is composed of within-household transmission $\beta_a^{hh}$ and between-household transmission $\beta_a^{bhh}$, such that $\beta_a^{hh} = \frac{\sigma S(\eta - 1)}{R_0}\beta_a$ and $\beta_a^{bhh} = \left(1 - \frac{\sigma(\eta - 1)}{R_0}\right)\beta_a$. | Estimated based on the equation for $R_0$, $f^{at}$ and $f^{pt}$ |
| $\beta_p$ | 0.54 | Daily transmission rate of pre-symptomatic cases. $\beta_p^{hh} = \frac{\sigma(\eta - 1)}{R_0}\beta_p$ and $\beta_p^{bhh} = \left(1 - \frac{\sigma(\eta - 1)}{R_0}\right)\beta_p$. | Estimated based on the equation for $R_0$, $f^{at}$ and $f^{pt}$ |
| $\beta_s$ | 0.52 | Daily transmission rate of symptomatic cases. | Estimated based on the equation for $R_0$, $f^{at}$ and $f^{pt}$ |

| | | | |
|---|---|---|---|
| | | $\beta_s^{hh} = \frac{\sigma(\eta-1)}{R_0}\beta_s$ and $\beta_s^{bhh} = \left(1-\frac{\sigma(\eta-1)}{R_0}\right)\beta_s$. | |
| $d^E$ | 3.8 (3.0, 6.2) | Mean duration of latent period (days). | Calculated by subtracting the mean pre-symptomatic duration $d^p$ [11] from a mean incubation period of 5.8 (5.01, 6.69) days [12] |
| $d^a$ | 7.7 (4.9,10.4) | Mean duration of asymptomatic infection (days) | [11], [13] |
| $d^p$ | 2 (0.5-4) | Mean duration of pre-symptomatic infection (days) | [11] |
| $d^s$ | 7 (4-16) | Mean duration of symptomatic infection (days) | [11] There is a large amount of variation in estimates of this parameter. It appears that, while patients may test positive for COVID-19 for considerably longer, the majority of transmission occurs within a week of symptoms onset. The duration of the symptomatic infectious period is also estimated to be lower in studies that consider children in addition to adults, and mild cases in addition to severe ones. We therefore anticipate that, for the purposes of this study, this parameter likely lies in the lower half of the possible range considered. |
| $d^{H_{wait}}$ | 7 (3-17) | Mean delay from symptoms onset to hospitalisation (general/ICU) (days) | [14], [15] |
| $d^{D_{wait}}$ | 20.2 (15.1, 29.5) | Mean delay from symptoms onset to death (days) | [14], [16] |
| $d^H$ | 5 (4-29) | Mean duration of stay in general hospital bed (days) | The selected value is the median total length of hospital stay estimated by [17] by combining estimates from a number of studies outside of China. The possible range is chosen to include the medians of all studies reviewed by [17], including those from China. |
| $d^{ICU}$ | 7 (5-19) | Mean duration of stay in ICU bed (days) | The selected value is the median ICU stay estimated by [17] by combining estimates from a number of studies outside of China. The possible range is chosen to include the medians of all studies reviewed by [17], including those from China. |

| | | | |
|---|---|---|---|
| $d^{Recup}$ | 21 | Mean duration of post-hospital discharge recuperation period (days) | 60% of ICU patients and 15% of general ward patients remained off sick from work when followed up a mean of 48 (range: 17-71) days post-discharge in the UK [18]. An additional 10% of ICU patients had switched from full-time to part-time work. |
| $d^{hh}$ | 10.56 | Mean duration for which households remain infected following introduction of the first infectious case | A stochastic version of the SEIR dynamics was run 100,000 times within a household of size $\eta$ with the within-household transmission rates from the calibrated model using the ssar package in R [1]. The mean time at which no infection remained in the household over all the simulations was calculated. |
| $d^{hha}$ | 9.87 | Mean duration for which households remain infected following introduction of the first infectious case, when that first case is asymptomatic | Estimated using the appropriate subset of the simulations used to estimate $d^{hh}$ |
| $d^{hhs}$ | 12.16 | Mean duration for which households remain infected following introduction of the first infectious case, when that first case is symptomatic | Estimated using the appropriate subset of the simulations used to estimate $d^{hh}$ |
| $f^{pt}$ | 0.23 (0.12-0.28) | Proportion of transmission by symptomatic cases that occurs in the pre-symptomatic period | These values are obtained by [19] using only cases that were not isolated before their 6$^{th}$ symptomatic day. Higher proportions are obtained when data are obtained under active case finding and isolation interventions [19], [20] |
| $f^{at}$ | 0.65 (0.20,1.25) | Multiplier of symptomatic transmission achieved by asymptomatic cases in the absence of intervention | The proportion of contacts of a group of symptomatic and asymptomatic infectious individuals that developed COVID-19 is reported by [21]. The value of the multiplier is obtained by dividing the proportion of asymptomatic contacts that developed COVID-19 by the proportion for symptomatic contacts. |
| $f^{a}$ | 0.701 | Proportion of cases that are asymptomatic | Population average calculated from the age-dependent values (Table S2), and the age distribution of the Dhaka District population from the 2011 census (Table S1) |

| | | | |
|---|---|---|---|
| $f^{H}$ | 0.073 | Proportion of symptomatic cases that are hospitalised | Population average calculated from the age-dependent values (Table S2), and the age distribution of the Dhaka District population from the 2011 census (Table S1) |
| $f^{ICU}$ | 0.31 | Proportion of hospitalised cases that require ICU | [22] |
| $f^{D}$ | 0.009 | Proportion of symptomatic cases that die | Population average calculated from the age-dependent values(Table S2), and the age distribution of the Dhaka District population from the 2011 census (Table S1) |
| $\sigma$ | 0.166 (0.140-0.193) | Probability with which an infected individual transmits to each of their household contacts (household secondary attack rate). | [2] |
| $t^{ld1}, t^{ld2}$ | 26/03/2020, 01/06/2020 | Start and end dates of the lockdown period. | Chosen to match the dates of the lockdown imposed by the government of Bangladesh |
| $f^{ldw}$ | 0.134 | Proportion of the population that is allowed to work throughout the lockdown, unless symptomatic. | The proportion of the employed population that is made up of key workers is assumed to match the value of 0.326 from the UK [23]. This is then multiplied by the proportion of the population that is employed $f^{e}$ (see below). |
| $\hat{\chi}^{ld}$ | 0.80 | Peak proportion of the population that is not included in $f^{ldw}$ that complies with the lockdown. | Assumption |
| $\varepsilon^{ld}$ | 0.97 | Proportion by which between household transmission drops during lockdown for people that are compliant with the lockdown. | Tuned to match ECDC (European Centre for Disease Prevention and Control) data for Bangladesh [8]. |
| $\tau^{ld}$ | 7 | Days after $t^{ld1}$ until the full effect of lockdown is reached. Compliance with the lockdown increases linearly from zero to $\hat{\chi}^{ld}$ during this period. | Assumption |
| $r^{ld}$ | 0.01 | Describes the rate of decline in lockdown compliance with time, after the improvement phase (see equation (S.6)). The changing pattern of compliance over time under the default parameters is illustrated in Fig. S1 | Assumption. Additional parts of the population, such as garment factory workers, were permitted to start returning to work mid-lockdown. Protests against the lockdown also began in April. Both of these factors led |

| | | | |
|---|---|---|---|
| | | | to a reduction in compliance over time. |
| $\hat{\chi}^{ld}_{\min}$ | 0.3 | Describes the minimum to which lockdown compliance can fall during the decline phase (see equation (S.6)) | Assumption |
| $t^{m1}, t^{m2}$ | 25/05/2020, 01/01/2022 | Start and end dates of the compulsory mask wearing period. | Default parameters in scenarios that include mask wearing assume that this intervention starts a week before the day lockdown ends and continues to the end of the modelled period. Sensitivity to this start date is considered within this study. |
| $\tau^{m}$ | 7 | Days after $t^{m1}$ until full effect of mask wearing is reached. Compliance with mask wearing increases linearly from zero to $\hat{\chi}^{m}$ during this period. | Assumption. There is expected to be some delay in reaching full effectiveness as people must access a supply of masks and learn to keep them on their person/use them most effectively. |
| $\hat{\chi}^{m}$ | 0.80 | Proportion of the population that complies with mask wearing. | Assumption |
| $\varepsilon^{m}$ | {0.2,0.5,0.8} | Proportion by which masks lower the wearer's virus emissions outside of the household. | The filtration efficiency of cloth masks varies widely depending on the material used, the number of layers, and the mask's quality of fit [24], [25]. The materials used in cloth masks typically have filtration efficiencies ≥50% [25], but poor fit qualities and failure of even compliant wearers to use their masks correctly may reduce this efficiency. Here, we consider three values of $\varepsilon^{m}$ representing low, medium, and high mask effects. |
| $\rho^{m}$ | {0,0.5,1.0} | The proportion by which masks lower the exposure of wearers to the virus outside of the household is taken to be $f^{mp}\varepsilon^{m}$. | As well as protecting others from transmission by the wearer, masks also provide some protection to the wearer themselves [26]. The protection to the wearer is assumed to be $\leq \varepsilon^{m}$ since use of masks as PPE requires them to block smaller particles than as source control [24]. We consider a range of fractions of $\varepsilon^{m}$ for the protective effect of masks. |
| $t^{q1}, t^{q2}$ | 25/05/2020, 01/01/2022 | Start and end dates of the community surveillance period | Default parameters in scenarios that include community surveillance assume that this intervention |

| | | | |
|---|---|---|---|
| | | | starts a week before the day lockdown ends and continues to the end of the modelled period. Sensitivity to this start date is considered within this study. |
| $\tau^q$ | 7 | Days after $t^{c1}$ until the full effect of community surveillance is reached. Household compliance with quarantine increases linearly from zero to $\hat{\chi}^c$ during this period. | Assumption. There is expected to be some delay in reaching full effectiveness as community health workers are mobilised and the general population starts to receive information about what symptoms should be reported and what the quarantine requirements are |
| $\hat{\chi}^q$ | 0.8 | Proportion of households in which a symptomatic individual occurs that are reached by and comply with messaging from community health workers to quarantine their households. | Assumption |
| $d^q$ | 14 | Days for which households detected by community surveillance must quarantine | 14 days is the quarantine duration now being imposed in Bangladesh |
| $N$ | 13 770 200 | Population size of Dhaka district in 2020 | Estimated from the proportion of the Bangladesh population that was in Dhaka District at the time of the 2011 census [4], and the UN population estimate for Bangladesh in 2020 [27] |
| $f^e$ | 0.52 | The proportion of the population that is employed is used to estimate working days lost by people who are affected by infection or by the lockdown and quarantine interventions. | Estimated using data for Dhaka District from the 2011 population and housing census [4] |
| $f^c$ | 0.23 | The proportion of the population that is unemployed, but works within the household, e.g. with caring responsibilities. This is used in the estimation of working days lost. When an individual in this group is hospitalised, recuperating, or dies, an individual in the employed population may have to stop working to take over caring responsibilities. | Estimated using data on household workers in Dhaka District from the 2011 population and housing census [4] |
| $\eta$ | 4 | The average household size in Bangladesh. This is used to determine how many people are at risk of | [3] |

| | | | |
|---|---|---|---|
| | | transmission from an infectious individual within their household and need to quarantine alongside an individual that develops symptoms during the community surveillance period. It is also used to determine the number of people who lose working days to grieve when a household member dies of COVID-19. | |
| Hospital beds | 10 947 | The number of hospital beds in Dhaka is used to inform estimates of excess cases that should be hospitalised but cannot be due to lack of beds. | Total number of general hospital beds estimated from UN estimates of beds per 10,000 in Bangladesh (7.95 in 2016) [28]. |
| $g$ | 7 | Number of days the rest of the household take off work to grieve when another member dies of COVID-19 | Assumption |
| $\nu$ | 0.27 | Proportion of households where members contract a non-COVID-19 influenza-like illness (ILI) over the course of a year. Under community surveillance these households are assumed to also be instructed to quarantine, affecting the working days lost. | It is assumed that 35% of people contract an ILI each year [29]–[31], and that, of those, 23% are members of a household where another individual has had/will have an ILI within the quarantine period, based on secondary infection risk of 10% [32], [33] and the average household size. |

**Table S5: Estimates of $R_0$ for the 2021 COVID-19 resurgence in Dhaka District.** Estimates were obtained by optimising $R_0$ under nine different scenarios of initial infectious and immune by two different approaches: 1) minimising the difference between modelled and reported cumulative deaths over the period from 1st March-5th April 2021 (Match deaths), and 2) minimising the difference in the timing of the peak in reported cases that occurred in late March and the peak in modelled symptomatic cases (Match peaks). Based on simulations using the optimised $R_0$ values, we also report differences in modelled and reported deaths and in the timing of the peaks in daily reported cases and modelled symptomatic cases during the resurgence.

| Initial infectious | Immunity (%) | $R_0$ | | Reported COVID-19 deaths detected between 1st March and 20th May as a proportion of modelled deaths | | Date of peak in daily reported cases – Date of peak in daily modelled symptomatic cases (days) | |
|---|---|---|---|---|---|---|---|
| | | Match deaths | Match peak timing | Match deaths | Match peak timing | Match deaths | Match peak timing |
| 5726 | 0 | 3.10 | 4.70 | 0.28 | 0.08 | -4 | 0 |
| 11451 | 0 | 2.53 | 4.12 | 0.46 | 0.09 | -3 | 0 |
| 17177 | 0 | 2.19 | 3.90 | 0.63 | 0.09 | -3 | 0 |
| 5726 | 25 | 4.08 | 5.97 | 0.32 | 0.11 | -4 | 0 |
| 11451 | 25 | 3.32 | 5.30 | 0.5 | 0.12 | -3 | 0 |
| 17177 | 25 | 2.87 | 4.67 | 0.68 | 0.13 | -2 | 0 |
| 5726 | 50 | 6.06 | 8.20 | 0.38 | 0.18 | -3 | 0 |
| 11451 | 50 | 4.90 | 7.20 | 0.57 | 0.19 | -2 | 0 |
| 17177 | 50 | 4.22 | 6.23 | 0.75 | 0.22 | -2 | 0 |

**Supplement C: Supplementary Figures**

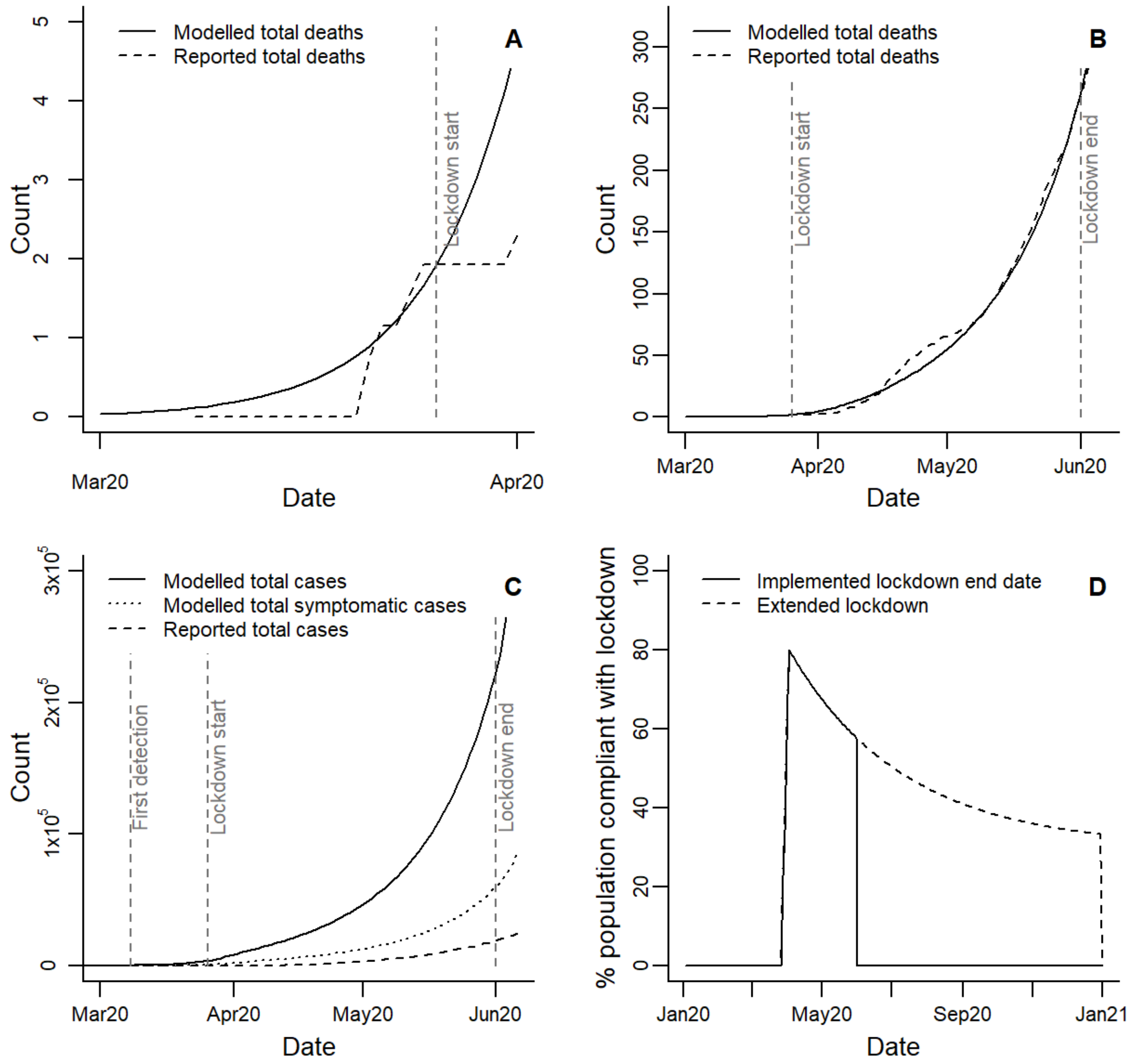


**Figure S1: Model calibration and parameterisation.** A) $R_0$ was optimised to minimise the absolute difference between modelled total deaths on the date lockdown started and the estimated total deaths in Dhaka district based on European Centre for Disease Prevention and Control (ECDC) data for Bangladesh (see *Methods*). This plot compares modelled to data-based deaths throughout the pre-lockdown period. B) The proportion of between-household transmission prevented for people complying with lockdown, $\varepsilon^{ld}$ , was optimised to minimise the difference between modelled deaths and ECDC data-based deaths on the date lockdown ended. This plot compares modelled to data-based deaths throughout the lockdown period. C) Comparison of the number of daily new cases (total and symptomatic-only) produced by the calibrated model and reported cases estimated from the ECDC data. D) The changing compliance of the population with lockdown based on the assumed parameters (supplementary Table S4). The solid line indicates the lockdown as implemented, which ended in June, while the dashed line indicates how we expect compliance to change under an extended lockdown scenario until the end of 2020.

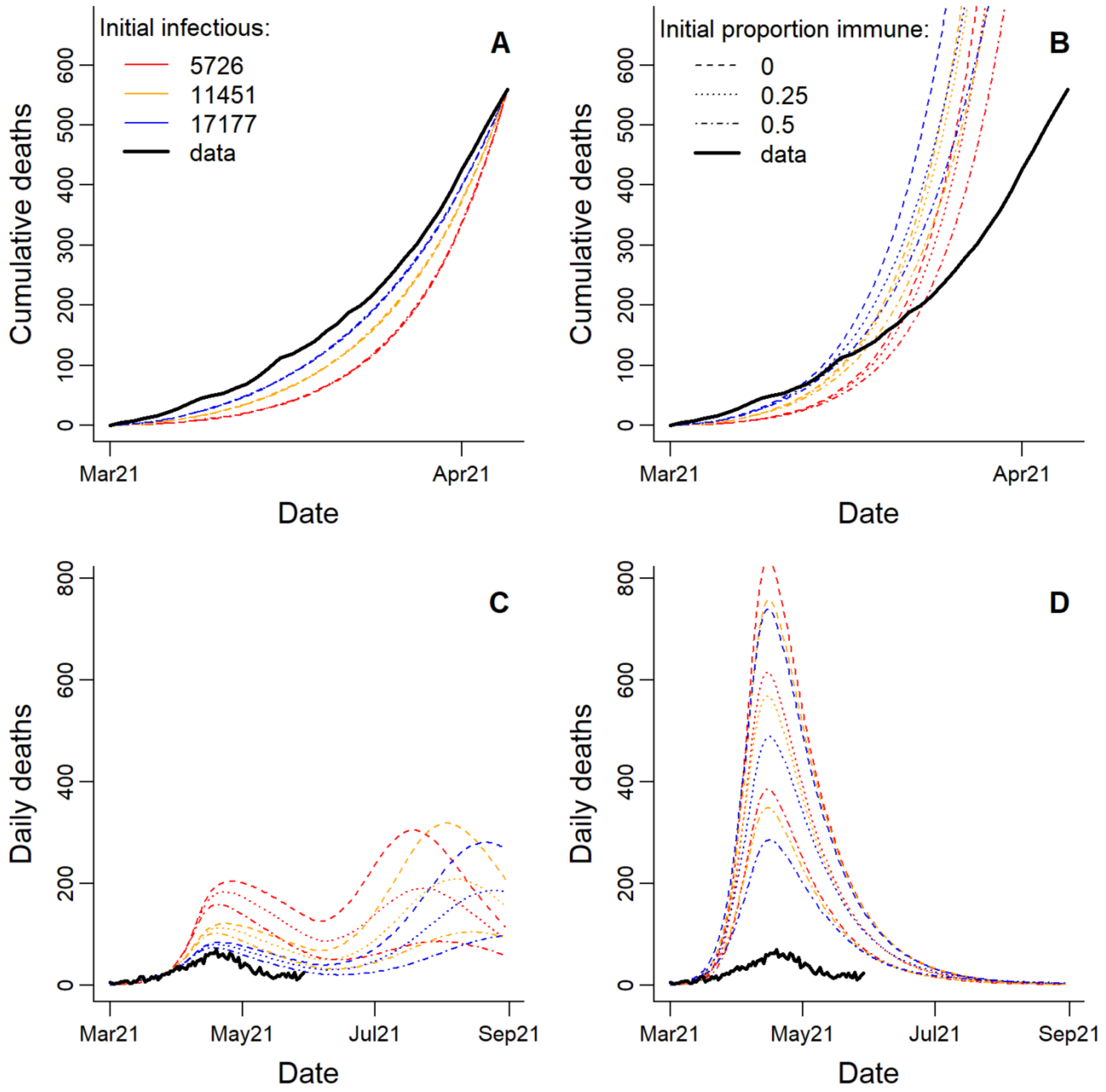


**Figure S2: Model calibration for 2021 resurgence.** When calibrating the model for 2021, $R_0$ was optimised under nine different scenarios of initial infectious and immune (see *Methods*) by two different approaches: 1) minimising the difference between modelled and reported cumulative deaths over the period from 1st March-5th April 2021, and 2) minimising the difference in the timing of the peak in reported cases that occurred in late March and the peak in modelled symptomatic cases. Plots A and C were obtained following optimisation by approach 1), and plots B and D were obtained following approach 2). A and B compare reported cumulative deaths to modelled cumulative deaths under each initialisation scenario over the first month of the resurgence. C and D compare reported daily deaths with modelled daily deaths under each initialisation scenario over an extended time period.

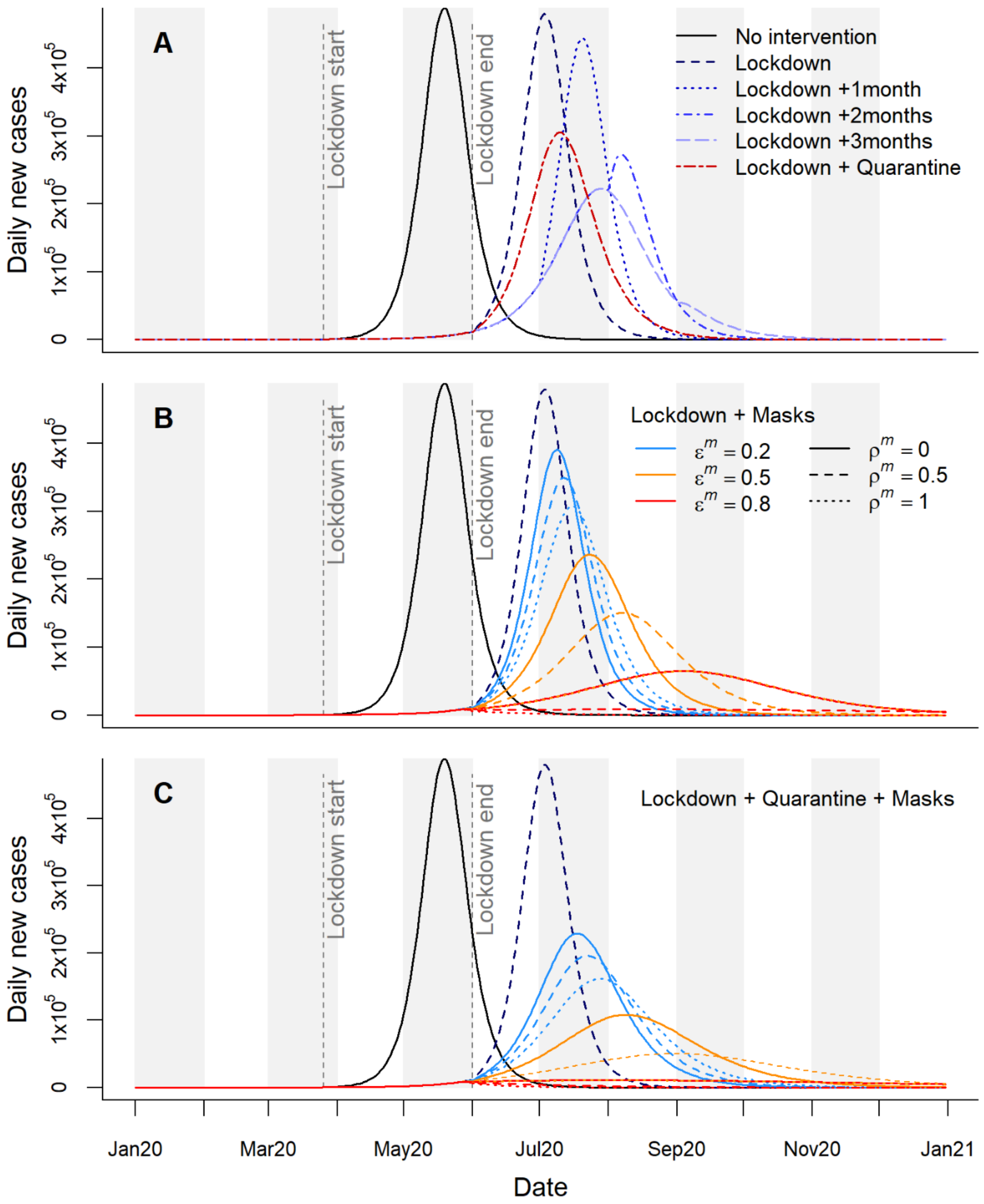


**Figure S3: Time series of daily new cases for different intervention scenarios.** Vertical lines indicate the start and end points of the lockdown as it was implemented in Bangladesh. A) Daily new cases in the absence of interventions, for the lockdown as implemented and with extensions of up to 3 months, and for the lockdown followed by household quarantine with community support teams. B) Lockdown as implemented followed by compulsory mask wearing, considering nine mask effectiveness scenarios; $\varepsilon^m$ describes the proportion reduction in outward emissions by mask wearers, while $\rho^m \varepsilon^m$ describes the proportion protection provided to mask wearers from others' emissions. C) Combined impacts of the lockdown, household quarantine, and masks of different effectiveness.

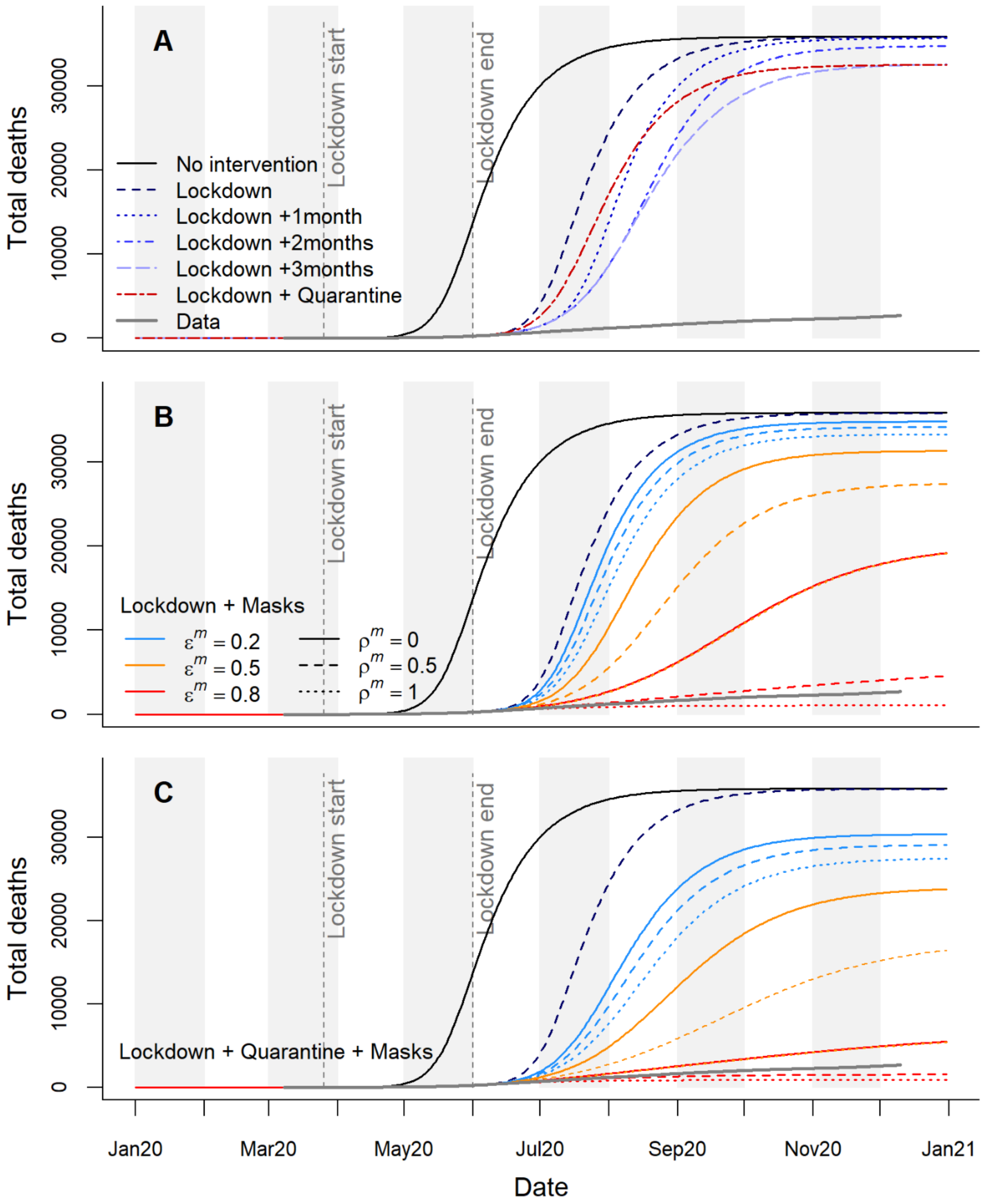


**Figure S4: Time series of cumulative deaths for different intervention scenarios.** The solid grey line shows the deaths in Dhaka District as estimated from the ECDC data. Vertical lines indicate the start and end points of the lockdown as it was implemented in Bangladesh. A) Modelled deaths in the absence of interventions, for the lockdown as implemented and with extensions of up to 3 months, and for the lockdown followed by household quarantine with community support teams. B) Lockdown as implemented followed by compulsory mask wearing, considering nine masks effectiveness scenarios; $\varepsilon^m$ describes the proportion reduction in outward emissions by mask wearers, while $\rho^m\varepsilon^m$ describes the proportion protection provided to mask wearers from others' emissions. C) Combined impacts of the lockdown, household quarantine, and masks of different effectiveness

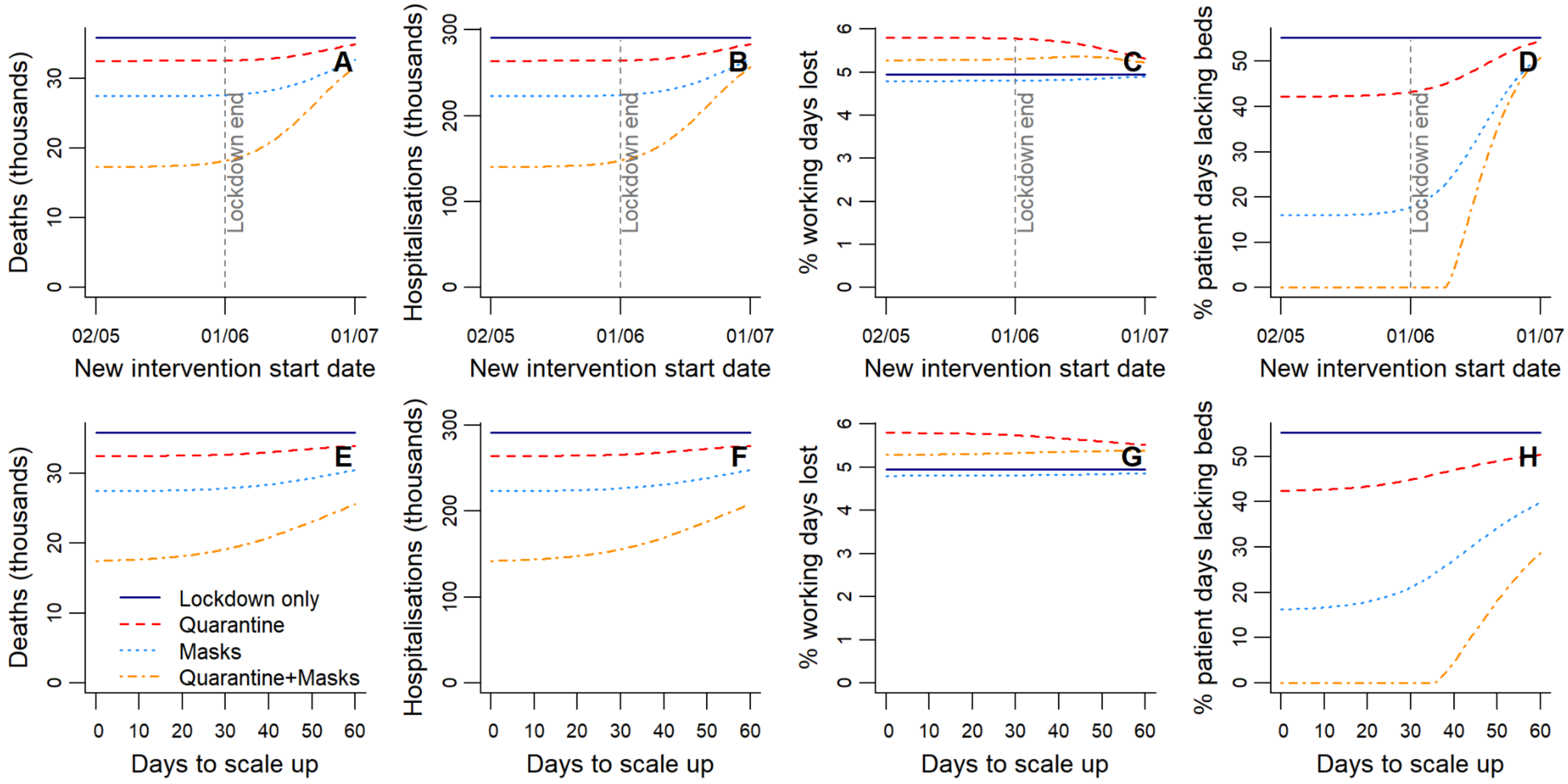


**Figure S5: Sensitivity to the start date and scale-up period of quarantine or mask wearing following lockdown.** A-D) Changes in health outcomes and working days lost calculated over 2020 and 2021 when the start date of interventions (household quarantine, masks, or quarantine and masks) following the initial lockdown is adjusted relative to the lockdown end date. The days taken to scale up post-lockdown interventions to their full effectiveness is held constant at seven days. E-H) Changes in the same outcomes when the days taken to scale up post-lockdown interventions is varied. The start date of the post-lockdown interventions is held constant at seven days prior to the lockdown end date.

**Supplement D: Sensitivity Analysis**

$R_0$ was among the top three most influential parameters for all five outcome measures reported under the baseline scenario (and also under the alternative baselines; Figs. S6-9). Reducing $R_0$ to the lowest value in the plausible range (1.56) had a large impact on cases, patients and deaths, slowing the epidemic to the point where it was still emerging in late 2020, while increasing $R_0$ had a much more limited impact. Under the baseline scenario with no interventions (Fig. S6), the next most influential parameter for total numbers of deaths, hospitalisations and cases was the household secondary attack rate, which gave a percentage change smaller of only around $\pm 0.1\%$. The duration of symptoms was the most influential parameter in determining working days lost (since symptomatic people are unable to work), followed by $R_0$, and, with a fairly small impact of less than $\pm 4\%$, the SARS-CoV-2 introduction date. Predictably, the mean lengths of stay in general hospital and ICU beds were highly influential for the percentage of patient days that lacked beds.

The sensitivity analysis using the lockdown as implemented scenario as a baseline produced results that were similar to the no intervention scenario, though the introduction date became more influential in determining outcomes (Fig. S7). When taking a scenario with lockdown followed by household quarantining as the baseline, the transmission rate of asymptomatic cases (as determined by $f^{at}$) emerges as a further important parameter in determining deaths, hospitalisations and cases (Fig. S8). This probably results from household quarantining mitigating transmission from symptomatic cases, making asymptomatic individuals more important in maintaining transmission. The duration of the symptomatic period also becomes less important for working days lost, possibly because working days lost are being more driven by the fixed duration quarantine period. When a scenario of lockdown followed by a period of mask wearing is used as the baseline for the sensitivity analysis, the length of the latent period becomes more influential on cases, hospitalisations, deaths, and working days lost (Fig. S9). This is probably because the slowing down of the epidemic provided by masks of moderate effectiveness, combined with the additional delaying effect of a longer latent period pushes the tail of the epidemic into 2021, reducing impacts in 2020.

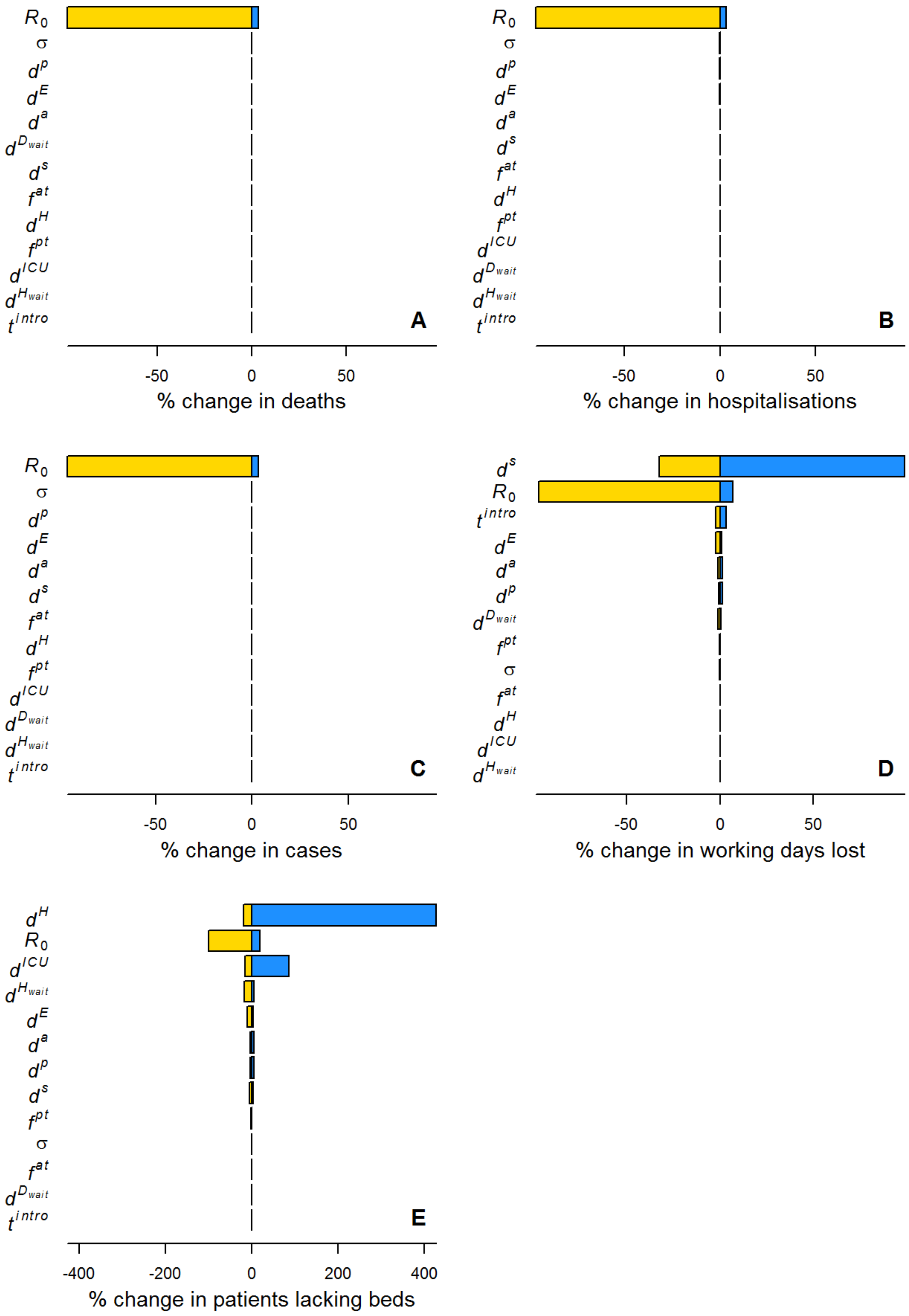


**Figure S6: Sensitivity analysis** on a range of epidemiological parameters (see Table S4 for ranges considered) using a baseline parameterisation with no interventions.

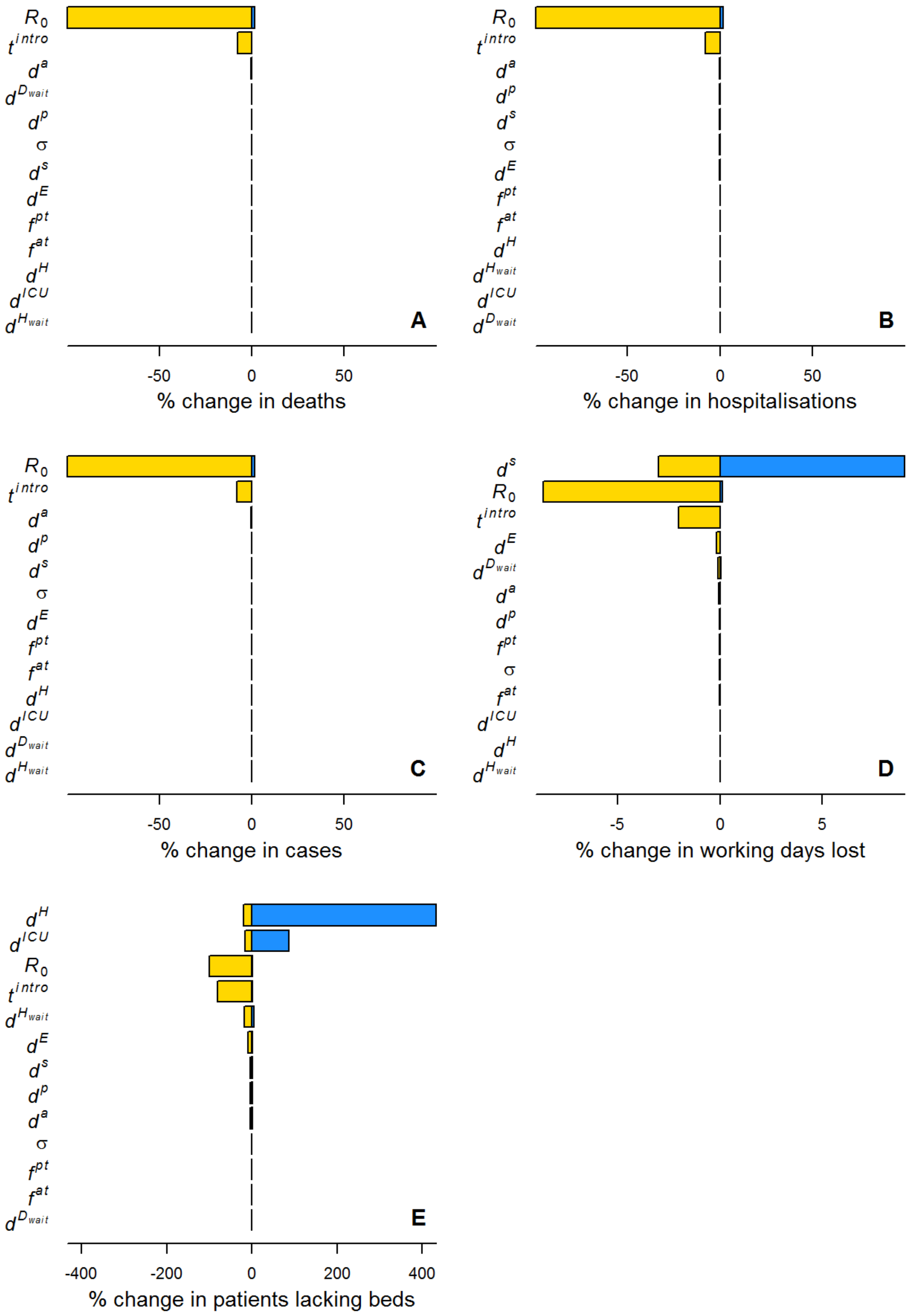


**Figure S7: Sensitivity analysis** on a range of epidemiological parameters (see Table S4 for ranges considered) using a baseline parameterisation with lockdown as implemented in Bangladesh.

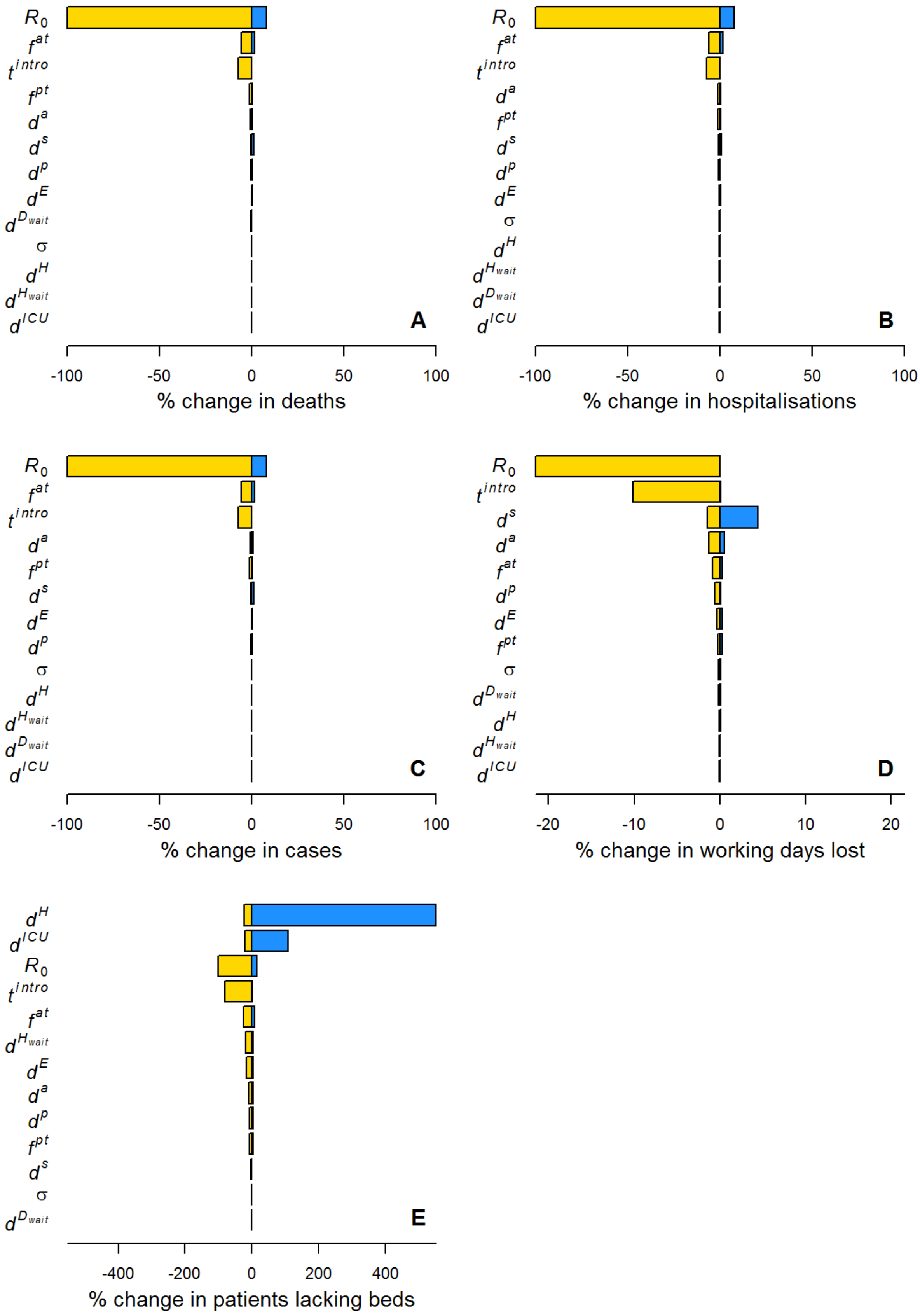


**Figure S8: Sensitivity analysis** on a range of epidemiological parameters (see Table S4 for ranges considered) using a baseline parameterisation with lockdown as implemented and household quarantine.

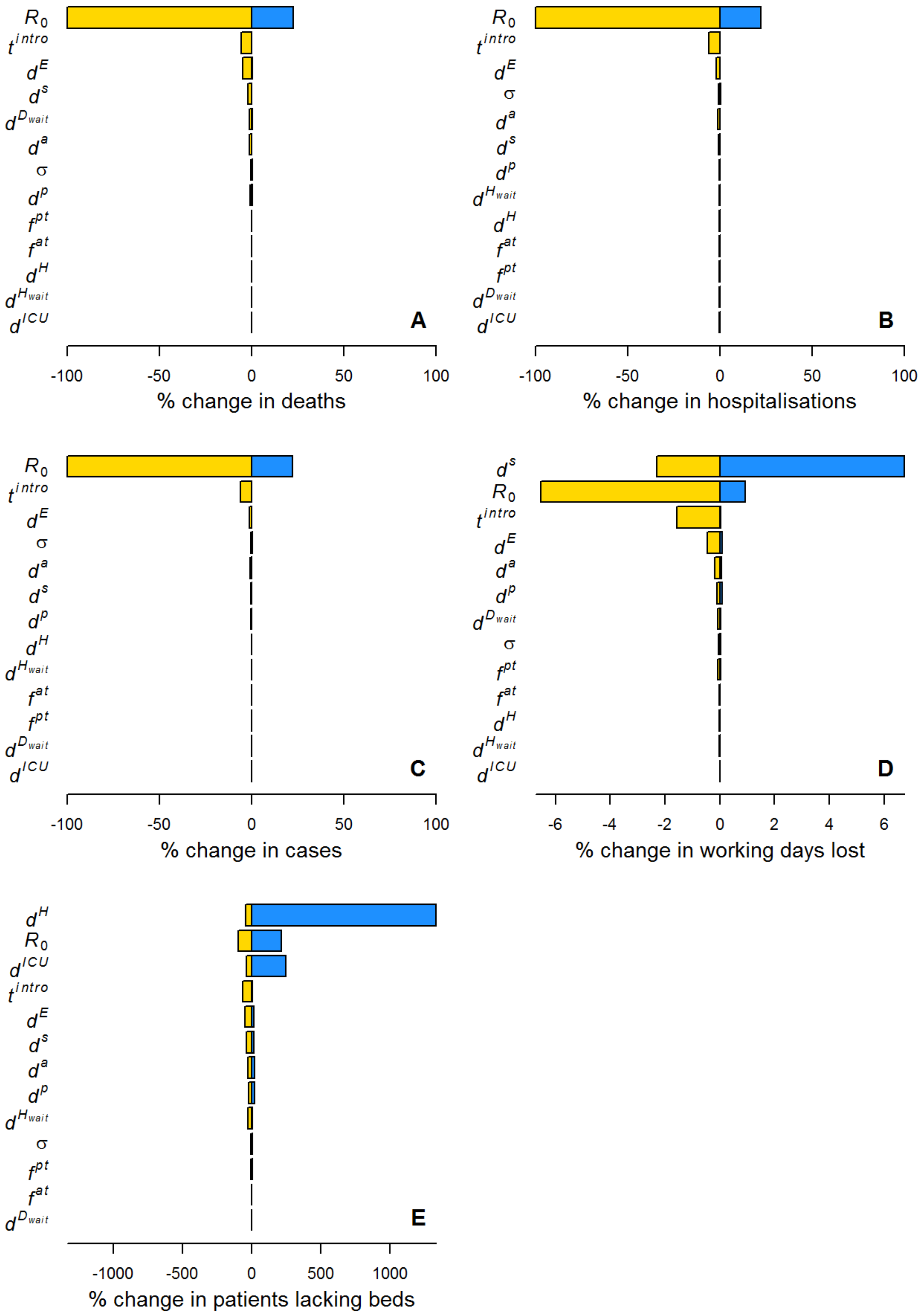


**Figure S9: Sensitivity analysis** on a range of epidemiological parameters (see Table S4 for ranges considered) using a baseline parameterisation with lockdown as implemented and mask wearing ( $\varepsilon^m = 0.5$ and $\rho = 0.5$ ).

[3] Bangladesh Bureau of Statistics, “Household Income and Expenditure Survey (HIES),” 2016.

[4] Bangladesh Bureau of Statistics, “Bangladesh Population and Housing Census 2011,” 2011. [Online]. Available: http://203.112.218.65:8008/Census.aspx?MenuKey=43.

[5] N. Davies *et al.*, “The impact of Coronavirus disease 2019 (COVID-19) on health systems and household resources in Africa and South Asia,” *medRxiv*, no. 0, 2020.

[6] N. G. Davies, P. Klepac, Y. Liu, K. Prem, M. Jit, and R. M. Eggo, “Age-dependent effects in the transmission and control of COVID-19 epidemics,” *Nat. Med.*, vol. 26, pp. 1205–1211, Aug. 2020.

[7] L. A. Cowley *et al.*, “Genomic and mobility data reveal mass population movement as a driver of SARS-CoV-2 dissemination and diversity in Bangladesh,” *medRxiv*, p. 2021.01.05.21249196, 2021.

[8] European Centre for Disease Prevention and Control, “Download today’s data on the geographic distribution of COVID-19 cases worldwide.,” 2020.

analysis," *Lancet Infect. Dis.*, vol. 20, no. 6, pp. 669–677, Jun. 2020.

[22] World Health Organisation (WHO), "Report of the WHO-China Joint Mission on Coronavirus Disease 2019 (COVID-19)," 2020.
